# Incorporating tandem repeats improves gene expression prediction models

**DOI:** 10.64898/2026.10.30.26366950

**Authors:** Michael Lamkin, Yang Li, Kai Akamatsu, Tiffany Amariuta, Melissa Gymrek

## Abstract

Tandem repeats (TRs), consisting of consecutive repeated units of 1+bp, are highly polymorphic features found throughout the genome that have been shown to have widespread impacts on gene expression and other traits. However, despite their prevalence, TRs are not included as features in polygenic prediction models, which typically focus on common single nucleotide polymorphisms (SNPs). Here, we develop a framework to incorporate TRs into gene expression models, which consider variants in a *cis*-window around each gene to predict expression levels. Simulation analyses demonstrate that including linear effects of TR copy number as features can result in substantially improved predictions, particularly when high impact causal TRs are poorly tagged by individual SNPs. We applied our framework to train and evaluate SNP+TR expression prediction models in lymphoblastoid cell lines (LCLs) using data from the Geuvadis cohort (n=359 European individuals) and compared these to SNP-only models. Overall, across 2,553 genes for which models trained successfully, we observed that SNP+TR models give significantly improved performance for 78 genes and a mean gain in adjusted R-squared 0.0037 across all genes, with most gains replicating in the independent Gene-Tissue Expression Project (GTEx) cohort. We additionally find that incorporating TRs results in simpler models and improves portability of models when testing on data for 89 Yoruban individuals from Geuvadis. Finally, we extended our models to consider nonlinear effects of TR copy number on expression. Under our nonlinear TR association testing, we identify 2,255 nonlinear significant genes not captured by linear association testing models, and identify exemplar genes, including *PALD1*, with highly polymorphic TRs for which incorporating their nonlinear effects results in substantially improved gene expression prediction. Overall, these results demonstrate the utility of including TRs in expression prediction models and show promise that they will have similar impact across other models such as polygenic risk scores for complex traits.

## Introduction

Genome-wide association studies (GWAS) have been instrumental in identifying genetic variants associated with complex traits. However, GWAS cannot directly discern how genetic variants in associated regions mechanistically impact a trait. Transcriptome-wide association studies (TWAS)^1–4^ attempt to address this shortcoming by identifying individual genes with genetically predicted expression levels that are associated with a trait such as diseases. Compared to GWAS, TWAS offers easier interpretability by focusing on genes rather than individual variants; has improved power due to reduced multiple testing burden; and has the potential to generate tissue-specific hypotheses^5^. TWAS typically first imputes expression values for each individual based on pre-trained linear models to predict tissue-specific gene expression from genetic variants^1,2,6^. Mostly, these gene expression prediction models focus on common single nucleotide polymorphisms (SNPs) and small indels in a *cis*-window around the gene. They have not typically considered the contribution of more complex variant types such as repeats and structural variants which have been shown in some cases to have large contributions to expression^7–11^.

Tandem repeats (TRs), which consist of repeating units of 1 or more bp in tandem, comprise over 2 million distinct loci throughout the human genome^12,13^. We^10,14^ and others^9,15,16^ have identified thousands of TRs at which variation in copy number is predicted to causally impact expression and we previously estimated that TRs explain around 10-15% of the *cis*-heritability of gene expression in humans^14^. Further, recent studies have identified hundreds of TRs predicted to be the causal drivers underlying top GWAS loci for a range of complex traits including blood traits^17^, height^7^, cancer risk^18^, and intellectual disability^19^. However, due to their rapid mutation rates and high multi-allelicness, TRs are often imperfectly tagged by nearby SNPs^20^ and thus their effects might not be well captured by SNP-based gene expression prediction models.

Here, we evaluate incorporating TRs alongside SNPs into gene expression prediction models. To this end we used genotyping and RNA sequencing data from the 1000 Genomes Project^21,22^ and Geuvadis^23^ for 359 European individuals to generate gene expression prediction models for 12,605 genes in lymphoblastoid cell lines (LCLs). We evaluate the improvement of model predictions with TRs compared to SNP-only models, perform replication analysis in LCL data available from the Genotype-Tissue Expression (GTEx) Project, and compare cross-ancestry portability of SNP-only vs. SNP+TR models. Finally, we assess the contribution of nonlinear effects of TR copy number on expression and incorporate these effects into gene expression prediction models. Our results demonstrate that the models that incorporate TRs in some cases result in dramatic prediction performance improvement as well as simplified and more portable models. Overall, our study demonstrates the benefit of considering a range of variant types for gene expression prediction models and suggests incorporating TRs will also benefit related applications such as polygenic risk scores for complex traits and diseases.

## Results

### A framework for integrating TRs into gene expression prediction models

We constructed a framework to generate gene expression prediction models for each gene based on *cis*-variants within 500kb of the gene boundaries (**Fig. 1a**). For this, we train LASSO regression models including either single nucleotide polymorphisms (SNPs) alone, or SNPs in combination with TRs. Due to potential variability of training LASSO models with cross-fold validation, especially in settings with low sample size, we train 20 separate models in identical ways for each gene and report the average results across models (adjusted R-squared based on 3-fold cross validation; **Methods**) for the SNP and SNP+TR models.

**Figure 1:**
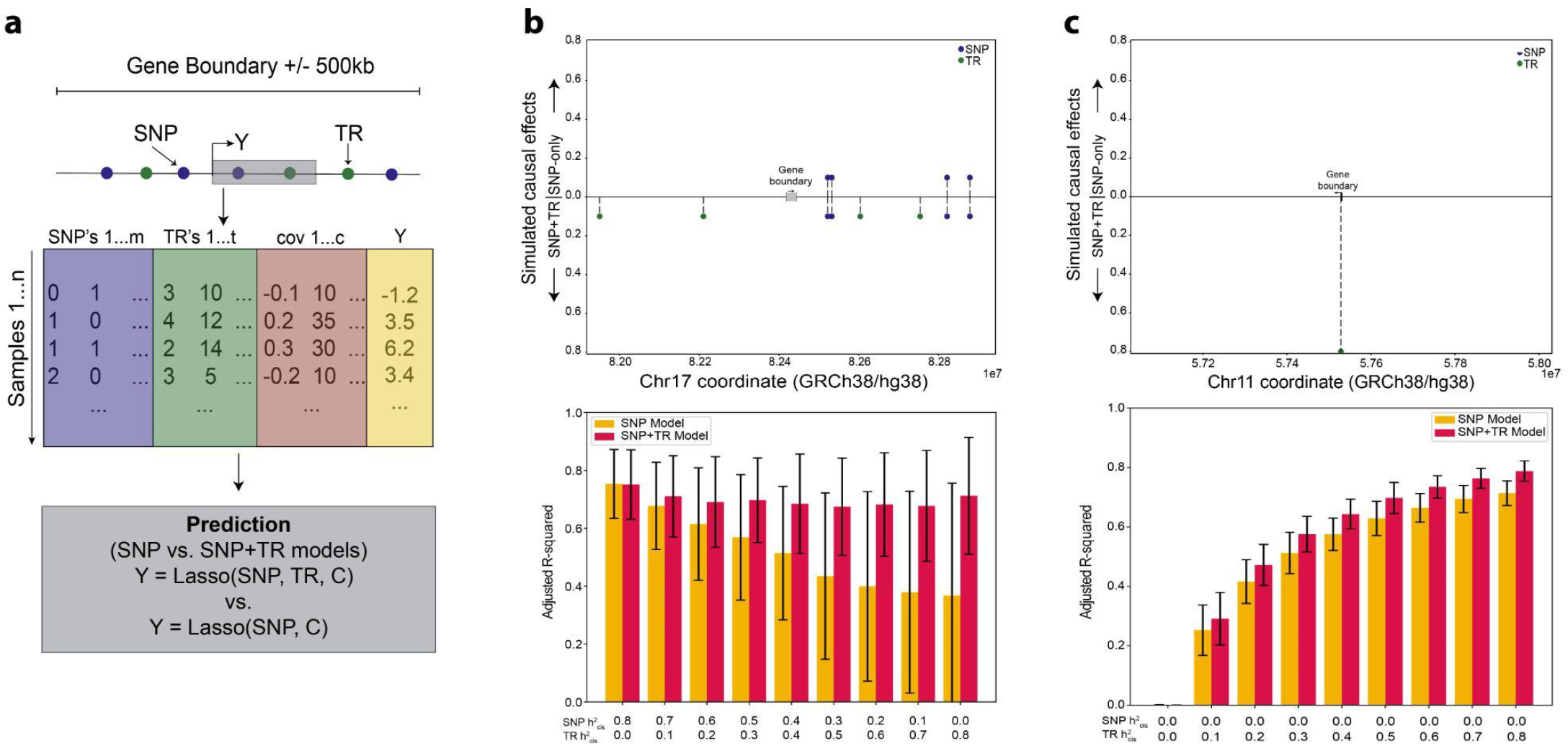
Integrating TRs into polygenic scores and simulation results. (a) Schematic overview of pipeline to train gene expression prediction models. We collect both SNPs and TRs within the gene boundary +/- 500kb as well as the top 5 PCs, sex, and 36 peer factors as covariates to train LASSO regression models for each gene. We then compare the resulting adjusted R-squared values from the models only trained on SNPs to the models trained on SNPs and TRs. (b) Evaluation of gene expression models on simulated expression based on multiple causal SNPs and TRs. In this setting we simulated up to four causal SNPs and TRs with a total *cis*-heritability of 0.8. The example is based on variants surrounding the gene *HEXDC*. (c) Evaluation of gene expression models on simulated expression based on a single causal TR. The example is based on variants surrounding the gene *TIMM10* with *cis-*heritability ranging from 0 to 0.8. For b-c, top panels show the set of causal SNPs and TRs included in each model, with y-axis values indicating SNP (blue) or TR (green) effect sizes (all were simulated to have positive effect sizes). The top half of the y-axis shows effects in the SNP-only setting and the bottom half shows the SNP+TR setting. Bottom panels show performance of the SNP only (gold) and SNP+TR (red) gene expression prediction models measured using adjusted R-squared based on 3-fold cross validation. The *cis*-heritability driven by SNPs vs. TRs is annotated for each setting on the x-axis. Error bars show 95% confidence intervals across 100 simulation rounds.

We first performed simulations to evaluate the robustness of our framework and gain insight into how the potential gain from including TRs in prediction models might vary across different genetic architectures of gene expression. Our simulations are based on real SNP+TR genotypes for 359 individuals of European (EUR) ancestry from the Geuvadis cohort, which are used in the real data analysis presented below. We evaluated performance under two different settings (**Fig. 1b-c**). In the first, we simulated a polygenic architecture with 8 total causal variants consisting of randomly chosen variants (4 SNPs and 4 TRs) with heritability explained distributed evenly across causal variants (**Methods**). The total heritability due to *cis*-variants was set to 0.8 and we varied the portion of *cis*-heritability (ℎ*_cis_*^2^ ) driven by SNPs vs. TRs from 0 to 0.8. In the second, we simulated a single causal effect for a highly polymorphic TR. The single causal effect matches patterns previously observed for the several strong eSTRs, including several known to influence medically relevant phenotypes (e.g. eSTRs for *CBL*^17^ and *NOP56*^10,24^). Simulations for this second setting are based on the *TIMM10* locus for which we previously identified a strong multi-allelic eSTR^25^. In this setting, we varied the total heritability explained by the TR from 0 to 0.8. For each setting, we repeated the simulation 100 times and reported the mean and standard error of the adjusted R-squared across simulations.

For the first setting (polygenic architecture with randomly chosen causal variants), we found that as expected SNP+TR models tend to outperform SNP-only models at genes for which a substantial portion of the *cis*-heritability of gene expression is driven by TRs (**Fig. 1b**). We observed similar results when varying the number of causal variants (**Supplementary Fig. 1**). In the second setting of a single large-effect highly polymorphic TR, we found that the SNP+TR models consistently generated models with adjusted R-squared close to the simulated *cis*-heritability (**Fig. 1c**), whereas the SNP-only models showed reduced performance (e.g. mean adjusted R-squared 0.788 for the SNP+TR model compared to 0.714 for the SNP-only model for ℎ*_cis_*^2^ =0.8). Across both simulation settings, we observed that gains for the SNP+TR models for a particular gene depend on how well causal TRs are tagged by nearby SNPs. In the first setting, simulations in which TRs were selected to have low LD with the best tag SNP show higher gains compared to simulations in which they are selected to have high LD with tag SNPs (**Supplementary Fig. 2-3**). Similarly, when repeating the simulation for the second setting using other causal TRs, we found that the gain in performance for the SNP+TR models expectedly reduces as the LD of the causal TR with the best tag SNP increases (**Supplementary Fig. 4**). Overall, our simulation results demonstrate that the subset of genes whose expression is driven by causal TRs that are not well tagged by *cis*-SNPs are expected to show substantial gains when incorporating TRs into gene expression prediction models.

### Constructing SNP+TR gene expression models in the Geuvadis LCL dataset

We applied the framework described above to construct both SNP-only and SNP+TR gene expression prediction models in the Geuvadis dataset, which includes RNA-sequencing from LCLs for 359 European (EUR; from 1000 Genomes^21^ populations CEU, FIN, GBR, TSI) and 89 Yoruban (YRI) individuals. SNP and TR genotypes have been previously generated for these samples based on available high-coverage whole genome sequencing^22,25^. After restricting to protein coding genes and filtering genes in segmental duplications or with low overall expression (**Methods**), 12,605 genes remained, of which 1,195 were estimated to have *cis*-heritability significantly different than zero, similar to previous estimates at similar sample sizes^1^. Gene expression values were adjusted for technical covariates and population structure prior to downstream analysis (**Methods**). After filtering on minor allele frequency (>0.01) and excluding variants failing Hardy Weinberg Equilibrium, 8,975,030 SNPs and 1,711,093 TRs remained for consideration in gene expression prediction models.

Of the 12,605 genes considered, LASSO models for either SNPs only or SNPs+TRs were successfully generated (non-negative adjusted R-squared) for 2,553 genes. Of these, we obtained SNP-only, SNP+TR models, or both, for 145, 203, and 2,205 genes, respectively. These genes are highly enriched for genes for which *cis-*heritability was estimated to be significantly non-zero (Fisher’s exact test odds ratio=21.57 one-sided *P*=1.35e-43; **Methods**). We evaluated model improvement using two metrics: the difference in adjusted R-squared (set to 0 when negative) based on 3-fold cross-validation and the difference in proportion of total *cis-*heritability explained between the SNP+TR vs. SNP-only models. Overall, SNP+TR models resulted in an average gain in adjusted R-squared of 0.0037 across all 2,553 genes analyzed. This gain was stronger for *cis*-heritable genes (0.0066; n=1,195) compared to non-heritable genes (0.0012; n=1,358; Mann Whitney [MW] U one-sided *P*=5.88e-5; **Methods; Fig. 2a**). The gain in adjusted R-squared exhibited a long tail, with 83 genes showing a gain of 0.05 or more (top 10 shown in **Table 1**) and 78 genes showing statistically significant gains (FDR<10%) in R-squared for SNP+TR models. We observed a similar trend when considering the percentage of *cis-*heritability explained (**Methods**), with on average an additional 1.79% explained by the SNP+TR vs. SNP-only models for the 2,553 genes but with a long tail of genes showing large improvements when adding TRs (**Supplementary Fig. 5**). Overall, our results show that incorporating TRs improves gene expression prediction models, with a subset of genes showing large predictive performance gains.

**Figure 2:**
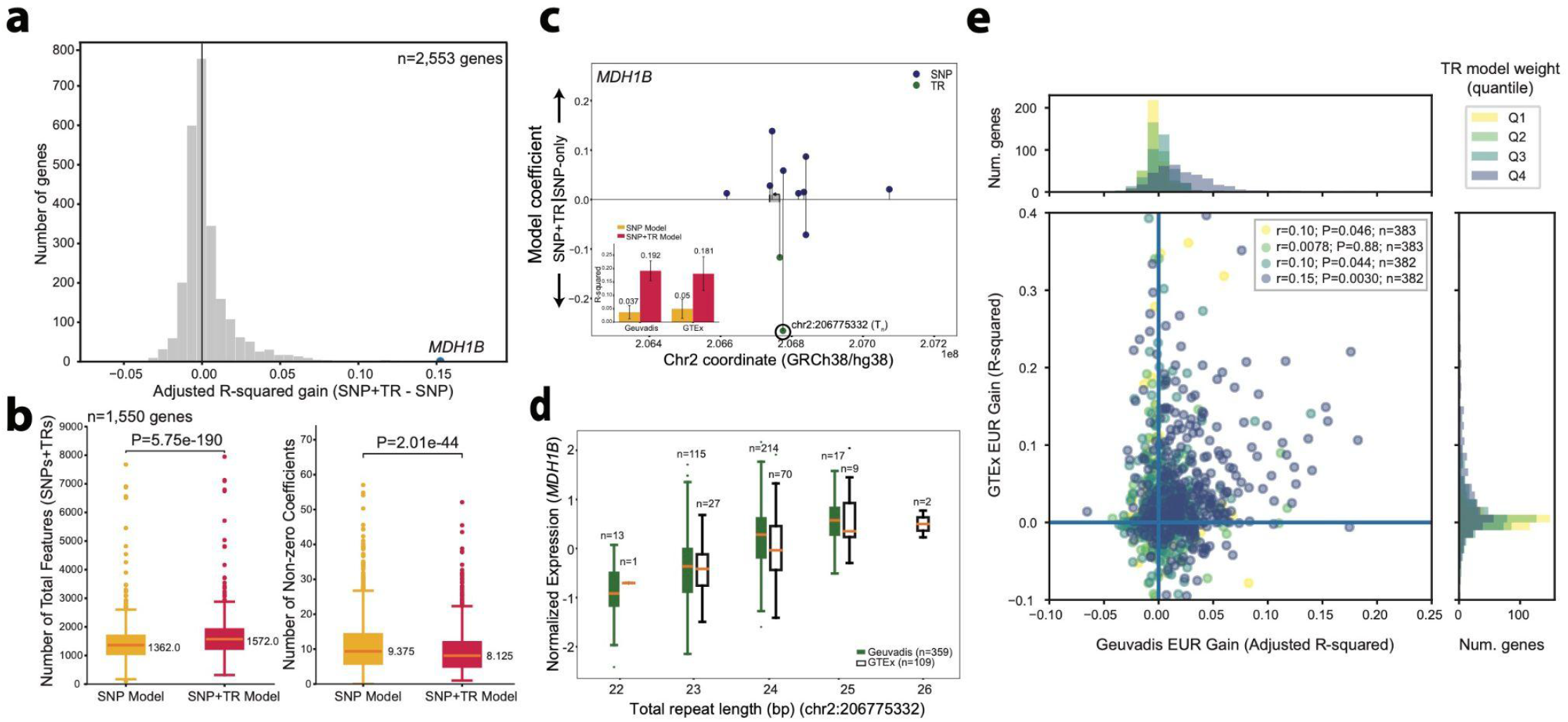
Evaluating SNP+TR vs. SNP-only gene expression models. (a) Distribution of the difference in adjusted R-squared for SNP+TR and SNP-only models across genes. Adjusted R-squared values are computed as the mean across 20 trained models for each gene for which at least one model had a non-zero adjusted R-squared (n=2,553 genes). The vertical black line denotes x=0. A positive value indicates the SNP+TR model had a larger mean adjusted R-squared (1,150 genes) and a negative value indicates the SNP model had a larger mean adjusted R-squared (1403 genes). **b) Comparison of the total number of input features and model sizes across SNP vs. SNP+TR models.** The left panel shows the distribution of the number of variants used to train models for each gene. The right panel shows the distribution of the number of variants that received non-zero coefficients (>1e-8) for each model. Values are averaged across all 20 models for each gene. For both panels gold=SNP-only models and red=SNP+TR models. (c) Model coefficients for *MDH1B* in SNP vs. SNP+TR models. The panel shows the absolute value of LASSO coefficients for each variant in SNP-only (top y-axis) and SNP+TR (bottom y-axis) models, based on the best performing model across all 20 trials. Blue=SNPs and green=TRs. The inset plot shows model performance of Geuvadis (adjusted R-squared) and GTEx (unadjusted R-squared) for SNP (gold) and SNP+TR (red) models. Error bars give 95% confidence intervals. (d) Association between the top TR and *MDH1B* gene expression. For the top TR (highlighted by a black circle in **(c)**), the plot shows the total repeat length (bp) across the two alleles of each individual (x-axis) vs. normalized *MDH1B* gene expression in Geuvadis (green) and GTEx (white). For **(b)** and **(d),** horizontal lines show median values, boxes span from the 25th percentile (Q1) to the 75th percentile (Q3), and whiskers extend to Q1-1.5*IQR (bottom) and Q3+1.5*IQR (top) where IQR is the interquartile range (Q3-Q1). **(e) Comparison of SNP+TR model gains across datasets**. We compared the gain in R-squared for SNP+TR vs. SNP-only models in the EUR cohort of Geuvadis (x-axis) vs. European samples in GTEx (y-axis) for each gene. Colors represent the quartile of each gene when ranked based on the total model weight assigned to TRs in trained models. Top and right panels show histograms of x-axis and y-axis values respectively stratified by these quartiles. The y-axis was truncated to remove several outlier data points (shown in **Supplementary** Fig. 12).

**Table 1.** Top 10 genes with largest gain in predictive performance for SNP+TR models vs. SNP-only models in the Geuvadis EUR cohort. Genes were ranked by the difference in average adjusted R-squared between the SNP+TR models vs. SNP-only models across the 20 trained models. When computing adjusted R-squared all negative values are converted to 0. Full results for all genes are available in **Supplementary** Table 1.

| Gene Name | Gene ID | EUR SNP Model Adjusted R Squared Mean | EUR SNP + TR Model Adjusted R Squared Mean | SNP Model #Nonzero Coeffs | SNP TR Model #Nonzero Coeffs |
|---|---|---|---|---|---|
| *FAM167A* | ENSG00000154319 | 0.220 | 0.402 | 46 | 21 |
| *RPL36AL* | ENSG00000165502 | 0.243 | 0.419 | 40 | 5 |
| *HLA-DPA1* | ENSG00000231389 | 0 | 0.175 | 54 | 10 |
| *MDH1B* | ENSG00000138400 | 0.037 | 0.192 | 10 | 3 |
| *SLC44A5* | ENSG00000137968 | 0.080 | 0.233 | 26 | 5 |
| *QDPR* | ENSG00000151552 | 0.105 | 0.245 | 8 | 5 |
| *SPATA20* | ENSG00000006282 | 0.379 | 0.519 | 27 | 14 |
| *NARF* | ENSG00000141562 | 0.061 | 0.198 | 34 | 10 |
| *MED16* | ENSG00000175221 | 0.137 | 0.263 | 23 | 7 |
| *MCM8* | ENSG00000125885 | 0.127 | 0.251 | 20 | 3 |

We hypothesized that in cases where the addition of TRs improved the model, it could be because one or more TRs contained predictive information that was either not captured by SNPs or required multiple SNPs to explain the effect of a single TR. To test this, we examined whether genes for which incorporating TRs was beneficial for prediction resulted in simpler overall models. This analysis was restricted to genes for which the difference of adjusted R-squared values between the SNP+TR model and SNP models was greater than 0, leaving a total of 1,150 genes. We quantified the complexity of trained models as the number of variants with non-zero LASSO coefficients (defined as the number of coefficients > ɛ averaged across the 20 models for each gene). Despite the increase in the number of features (variants) in the SNP+TR models (median 1,572 features) compared to the SNP-only models (median 1,362 features; Wilcoxon signed-rank test one-sided *P*=5.75e-190; **Fig 2b**), we saw that SNP+TR models had lower overall numbers of non-zero coefficients (median 8.125 with ɛ=10^-8^) compared to SNP-only models (median 9.375; Wilcoxon signed-rank test one-sided *P*=2.01e-44; **Fig. 2b**). These trends were robust to thresholds used to determine which genes showed improved adjusted R-squared or the value of ɛ used to determine the number of non-zero coefficients per model (**Supplementary Fig. 6**). Further, the reduction in the number of non-zero coefficients was strongest for genes with the highest improvement in adjusted R-squared (change in medians of 1.25 and 6.95 for gain in adjusted R-squared threshold of 0 and 0.05 respectively; **Supplementary Fig. 6**). Additionally, we found that at 1,009 genes for which SNP+TR models had positive gains in adjusted R-squared and contained nonzero TRs model coefficients, the performance gain was negatively correlated with the strength of the LD between the lead TR in the model and its best tagging SNP (Pearson *r*=-0.229, two-sided p-value=1.85e-13; **Supplementary Fig. 7).** This recapitulates our findings from simulations, in which TRs show the highest gain when high-impact TRs are in low LD with surrounding SNPs. An example gene, *MDH1B*, for which a SNP+TR model resulted in substantial performance gain and model simplification is shown in **Fig. 2c-d**. Incorporating TRs resulted in an adjusted R-squared of 0.1917 compared to 0.0368 for a SNP-only model. Further, the SNP-only model contained 8 non-zero coefficients (>10^-8^) compared to 2 TRs and 1 SNP for the SNP+TR model (**Fig. 2c**). The model gives by far the strongest weight to a homopolymer T_n_ repeat (start coordinate hg38:chr2:206,775,332) which shows a strong positive association with expression of *MDH1B* (**Fig. 2d**). Additional similar examples are shown in **Supplementary Fig. 8**. Overall, these results show that for a subset of genes multiple SNPs are needed to achieve the same predictive power as a single TR, suggesting the SNPs are acting as imperfect tags for TR effects.

To validate our models, we used our gene expression prediction models based on the Geuvadis cohort to predict gene expression in an orthogonal LCL dataset for 109 individuals predicted to have European ancestry (**Methods, Supplementary Fig. 9**) obtained from the GTEx project and compared to observed expression values. Although we previously generated genome-wide TR genotypes obtained from WGS for GTEx^10,26^, due to high missingness rates after quality filtering we instead relied on imputation to obtain TR genotypes using our published SNP+TR reference panel^25^. Of the 223,189 total TRs considered for gene expression prediction models in the Geuvadis analysis, 135,247 were in our reference panel and could be imputed into GTEx. We filtered genes that had no TRs included in the SNP+TR models (n=421 genes) and for which TRs included in the models were not in our imputation panel (n=602 genes), leaving 1,530 genes for the validation study. For these genes, only a subset of relevant TRs were present in the imputed data (**Supplementary Fig. 10**), and thus we stratified performance across genes based on the sum of absolute values of model weights of available TRs. Overall, we found the model R-squared values from GTEx to be highly concordant with adjusted R-squared values from Geuvadis (Pearson *r*=0.65; two-sided *P*=4.67e-185 for SNP models and *r*=0.68; *P*=7.34e-206 for SNP+TR models; **Supplementary Fig. 11**). Further, the gains in the SNP+TR vs. SNP-only models are also correlated across datasets (Pearson *r*=0.200; *P*=2.55e-15; **Supplementary Fig. 12**; **Fig. 2e**) with the strongest correlation for genes with the highest total weight assigned to TRs (Pearson *r*=0.15; two-sided *P*=0.0030; n=382 for genes with gains in the top quartile). For the example gene *MDH1B* shown in **Fig. 2c-d**, R-squared increases from 0.05 to 0.181 for SNP vs. SNP+TR models in GTEx, similar to observed values of 0.067 and 0.2 in Geuvadis, respectively. Overall, these results demonstrate the replicability of both SNP and SNP+TR models across independent datasets including the gain from including TRs. Additional similar examples are shown in **Supplementary Fig. 8**.

### Cross population improvement to prediction accuracy

We evaluated the ability of our gene expression prediction models trained on EUR samples to predict expression in individuals of Yoruban (YRI; Yoruba in Nigeria) ancestry. From the set of models trained above (20 models per gene per model type), we used the SNP and SNP+TR models that resulted in the largest adjusted R-squared to predict gene expression for 89 YRI samples with matched LCL expression data available in Geuvadis. Similar to the analyses above, we computed the Pearson *r^2^* between predicted and measured gene expression using either the SNP-only or SNP+TR models. Note we compute adjusted R-squared of the EUR data to account for overfitting since it was trained on EUR data, but compute unadjusted R-squared for the YRI samples as they are an independent dataset that was not used in model training. Overall, predictive performance of the EUR models across ancestries was highly correlated across genes (Pearson *r*=0.579, two-sided *P*=2.18e-228 and Pearson *r*=0.575, *P*=6.36e-225 for SNP-only and SNP+TR models, respectively; **Supplementary Fig. 13**). Similarly, the gains in R-squared in the SNP+TR vs. SNP-only models for each gene were correlated across the two cohorts (Pearson *r*=0.199, two-sided *P*=3.44e-24). YRI prediction results for each gene are provided in **Supplementary Table 1**.

Next, we sought to identify genes for which SNP+TR models showed the greatest gain over SNP-only models in the YRI vs. EUR cohorts. We hypothesized that genes for which the SNP+TR gain is larger in YRI individuals indicate examples where SNP models are sufficient to capture most variation including that driven by TRs in the training (EUR) cohort, but do not generalize well to other populations for which the model SNPs might not serve as adequate LD proxies for causal TRs. Overall, we observed that 1,423/2,553 (56%) of SNP+TR models showed higher gains in the YRI vs. EUR cohorts, with a median difference in gains of 0.0014 (**Fig. 3a**). For the 1,530 genes analyzed in the GTEx cohort, the difference in gains for each gene observed in Geuvadis for YRI vs. EUR samples was correlated with the difference in gains for AFR vs. EUR samples in GTEX (Pearson *r*=0.235; two-sided *P*=1.38e-20). Genes with the top differences in gain are shown in **Table 2**. The gene with the largest difference in gains, *TIMM10*, showed a gain in R-squared of 0.297 in the YRI cohort compared to only 0.038 in the EUR cohort, indicating that while TRs did not substantially improve predictions in EUR, their inclusion resulted in dramatic improvement in the YRI cohort (**Fig. 3b**). Inspection of model weights indicated that whereas the SNP-only model assigned modest weights (coefficients range 0.15-0.25) to four individual SNPs, the SNP+TR model assigned nearly all weight (coefficient=0.703) to a single highly polymorphic AT_n_ dinucleotide TR (hg38:chr11:57,528,484; **Fig. 3c**). This TR shows a strong linear association between total repeat unit length and normalized *TIMM10* expression levels in both cohorts (two-sided *P*=1.24e-114 for EUR and 4.34e-14 for YRI; **Fig. 3d**). We observed a similar trend in GTEx (**Fig. 3d**; **Table 2**), with no gain in R-squared for EUR (−0.026) compared to a gain of 0.24 in GTEx samples of African ancestry.

**Figure 3.**
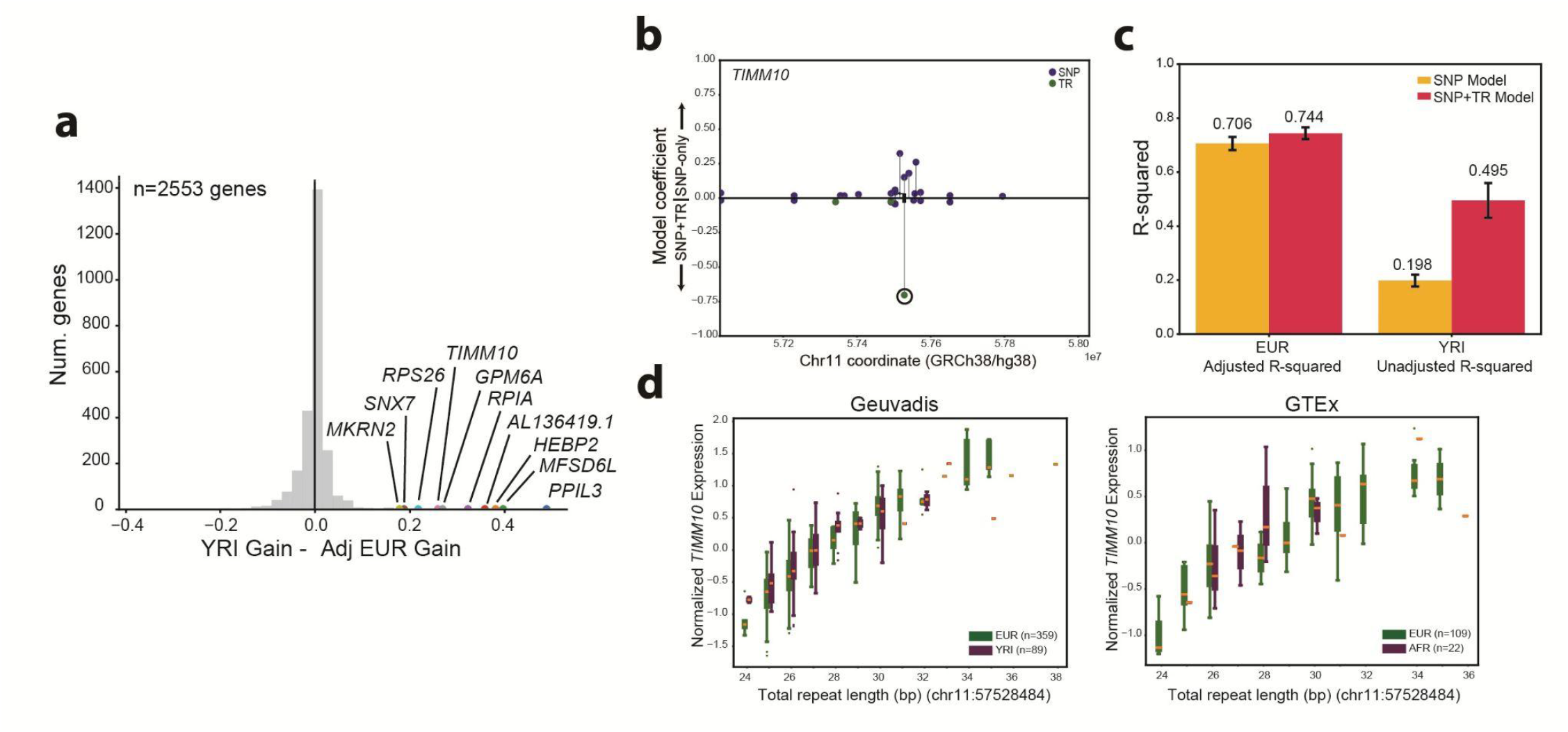
Evaluation of gene expression prediction models across ancestries. (a) Distribution of the difference in gain (SNP+TR-SNP model R-square) in YRI and EUR cohorts. The vertical black line gives x=0. The top 10 genes for YRI gain - adjusted EUR gain are highlighted. (b) Model coefficients for *TIMM10* in SNP vs. SNP+TR models. The panel shows the absolute value of LASSO coefficients for each variant in SNP-only (top y-axis) and SNP+TR (bottom y-axis) models, based on the best performing model across all 20 trials. Blue=SNPs and green=TRs. (c) Model performance in EUR vs. YRI samples for *TIMM10*. The y-axis gives R-squared values for EUR (adjusted R-squared) and YRI (unadjusted R-squared) for SNP (gold) and SNP+TR (red) models. Error bars give 95% confidence intervals. (d) Association between the top TR and *TIMM10* gene expression. For the top TR (highlighted by a black circle in **(b)**), the plot shows the total repeat length (bp) across the two alleles of each individual (x-axis) vs. normalized *TIMM10* gene expression in Geuvadis (left) and GTEx (right). Green=EUR samples and purple=YRI (Geuvadis) and AFR (GTEx) samples. Horizontal lines show median values, boxes span from the 25th percentile (Q1) to the 75th percentile (Q3), and whiskers extend to Q1-1.5*IQR (bottom) and Q3+1.5*IQR (top) where IQR is the interquartile range (Q3-Q1).

**Table 2.**
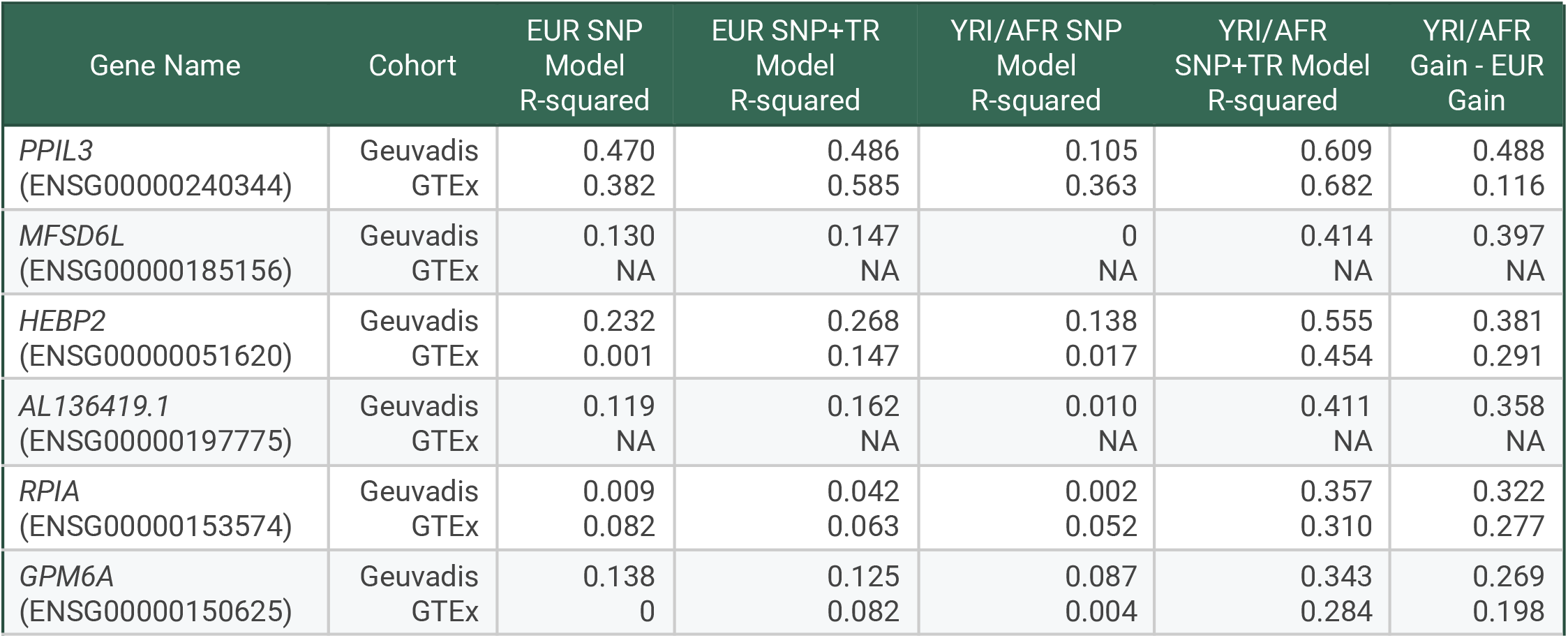
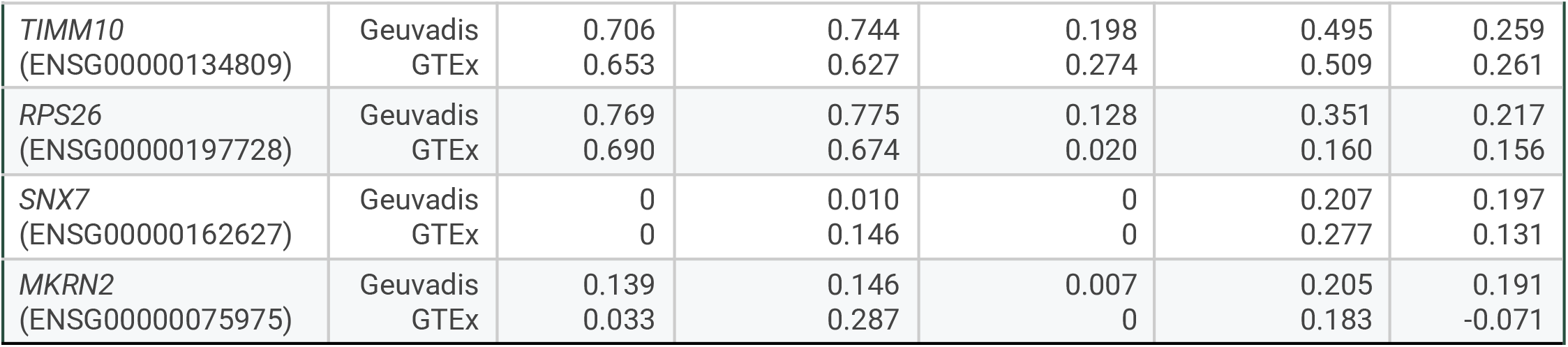
Top 10 genes with largest difference in model performance gains for SNP+TR vs. SNP models across populations. In each cohort, we computed gain as the difference in R-squared values for SNP+TR vs. SNP-only models. Genes are ranked by the difference in gains in YRI vs. EUR cohorts in Geuvadis. When computing adjusted R-squared all negative values are first converted to 0. In each cell, values computed by comparing EUR vs. YRI cohorts in Geuvadis are shown at the top followed by values computed by comparing EUR vs. AFR cohorts in GTEx at the bottom. For EUR Geuvadis, adjusted R-squared values are shown since that cohort was used in training. For other groups, unadjusted R-squared values are provided.

Taken together, these results suggest the TR is a causal driver of expression that is differentially tagged by common SNPs in different populations. As a result, a model incorporating TRs shows greater portability across the EUR and AFR cohorts. Additional example genes demonstrating similar gains in portability, but not necessarily EUR model performance, for SNP+TR compared to SNP-only models are shown in **Supplementary Fig. 14**.

The majority of both SNP (82%) and SNP+TR (84%) models showed stronger predictive performance in the EUR cohort in which they were trained. However, we observed a subset of SNP+TR models (n=455 genes) for which models showed stronger performance in the YRI cohort despite being trained on EUR. Of these, 114 genes did not have any TRs with non-zero coefficients and showed minimal improvements (average R-squared increase 0.0060 in YRI SNP+TR vs. SNP-only models) and thus likely do not represent meaningful differences. We further inspected the remaining 341 genes which contained at least one TR with a non-zero coefficient in the SNP+TR models. Of these, 180 genes had at least one TR with a non-zero model coefficient for which the range of alleles was greater in the YRI vs. EUR cohorts. One such example gene is *MFSD6L*, which shows an adjusted R-squared of 0.147 in the EUR cohort compared to an R-squared of 0.414 in the YRI cohort. The SNP+TR model for this gene contains 3 TRs with non-zero coefficients. Of these, two show similar allele frequency distributions across YRI and EUR samples. The third, a homopolymer A_n_ repeat at hg38:chr17:8802414, which has the largest absolute coefficient size (−0.31), shows a substantially wider range of TR lengths (26-47 copies) in YRI compared to EUR (36-48 copies) individuals but with a similar linear relationship between repeat length and *MFSD6L* expression as was observed in EUR (**Supplementary Figure 15**). Overall, this suggests that at a subset of genes for which SNP+TR models accurately capture TR associations with expression, models show outsized performance gains in populations for which those TRs have larger ranges of alleles and therefore might capture greater variance in expression than in the training cohort.

### Incorporating nonlinear effects of TRs on gene expression

The gene expression models described above consider only linear effects of TR copy number on expression. Given the existence of multiple known examples of nonlinear TR-phenotype associations in both humans and other species^27–29^, as well as our recent observations of widespread nonlinear effects of TRs on expression using a massively parallel reporter assay^28^, we evaluated whether modeling nonlinear effects of TR copy number on expression would improve our gene expression prediction models. Although multiple classes of nonlinear models exist, we focused on a quadratic model (**Methods**) given its relatively low model complexity (one additional parameter compared to linear models), limitations of sample sizes of existing expression datasets to fit more complex models, and the similar ability of a quadratic model to capture many types of effects (e.g. threshold, bell-shape curve) observed in real data compared to more complex alternatives.

We first sought to identify examples of nonlinear effects and quantify their prevalence in the LCL datasets of interest. For this, we fit both linear and quadratic models (**Methods**) to identify associations between TR copy number and expression of nearby genes (within the gene body +/- 500kb) in the 359 EUR samples from the Geuvadis cohort (**Fig. 4a**) and evaluated replication of effects in 109 EUR samples from GTEx. In total we performed 3,427,940 variant-gene association tests for each model across 12,605 unique genes. We identified 88,809 and 100,674 gene-TR pairs (3,266 and 3,339 unique genes) meeting a gene-level FDR<5% when applying the linear and quadratic models, respectively, with the linear association result in line with the total number of eGenes expected at this sample size based on previous studies^10,30^. Association statistics (−log_10_ P-values) for linear tests were strongly correlated between Geuvadis and GTEx (Pearson *r*=0.751; two-sided *P*<1e-200; n=16,961 TR-gene pairs with adjusted P<5% in Geuvadis that achieved nominal *P*<5% in GTEx; **Fig. 4b**). For 15,040 gene-TR pairs (2,255 unique genes), the quadratic model significantly improved the fit (one-way ANOVA adjusted P<0.05) compared to a linear model. Of these, 252 gene-TR pairs did not show significant linear associations and were only detected by nonlinear models. Nonlinear association statistics were strongly correlated between Geuvadis and GTEx, although to a lesser extent than linear association statistics (Pearson *r*=0.53; two-sided *P*<1e-200; n=18,545 TR-gene pairs with adjusted *P*<5% in Geuvadis that achieved nominal *P*<5% in GTEx; **Fig. 4c**).

**Figure 4.**
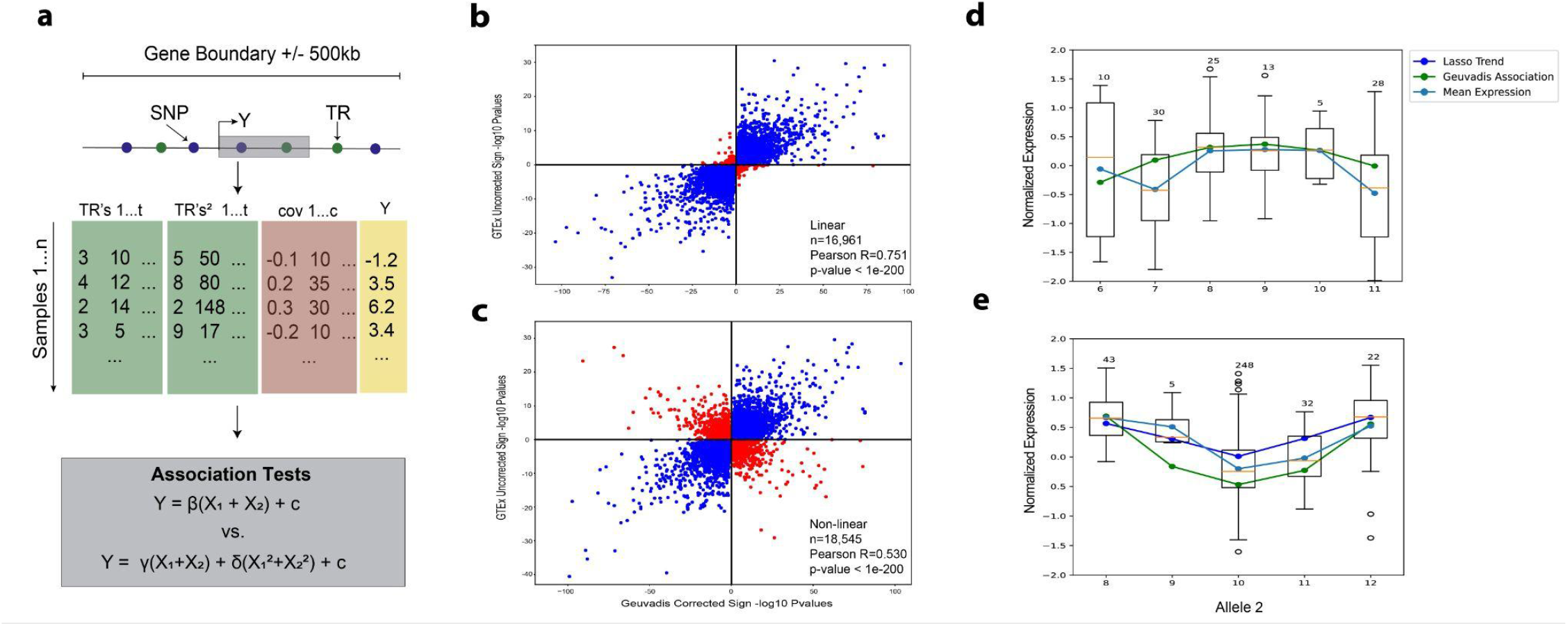
Evaluation of nonlinear effects of TRs on gene expression. (a) Schematic overview of pipeline to detect linear and nonlinear associations between TR copy number and expression of nearby genes. For the nonlinear model, we add an additional term consisting of the sum of squared repeat lengths to capture quadratic associations (**Methods**). **(b) Comparison of association statistics for linear models in Geuvadis vs. GTEx.** The panel shows signed −log_10_ P-values (based on the effect size direction) for effects with adjusted *P*<5% in Geuvadis and nominal *P*<5% in GTEx. Pearson *r*=0.751; two-sided *P*<1e-200; n=16,961 gene-TR pairs. **(c) Comparison of association statistics for nonlinear terms in Geuvadis vs. GTEx.** The panel shows the −log_10_ P-values for the quadratic coefficient (γ) for effects with adjusted P<5% in Geuvadis and nominal *P*<5% in GTEx. Pearson *r*=0.53; two-sided *P*<1e-200; n=18,545 gene-TR pairs. For **b-c**, blue=points with concordant effect size directions and red=points with discordant directions. **(d-e) Stratified associations with expression for example TRs with significant nonlinear associations.** For **(d),** the top panel shows the association between CCG_n_ copy number at hg38:chr3:19947266 and Z-normalized *EFHB* expression for all individuals with diploid genotypes of the form (6,X), where the x-axis value gives the value of X. Thus, only the second allele varies between individuals enabling visualization of the nonlinear effect. Blue=observed mean expression; green=trend learned from nonlinear association testing. Panel **(e)** is the same as **(d)** but shows data for a C_n_ at hg38:chr10:70,473,982 that is significantly associated with the gene *PALD1* and includes individuals with genotypes of the form (10, X). For **(e)**, dark blue = trend inferred from LASSO model coefficients at this locus. For **d-e**, the allele to hold constant (6 or 10 respectively) was chosen based on the most common allele at each locus. Additional plots based on holding other alleles constant are shown in **Supplementary** Fig. 15**-16**.

TRs passing ANOVA (n=12,707 unique TRs), meaning nonlinear models improve model fit compared to linear models, are significantly more heterozygous (Mann-Whitney two-sided *P*<1e-200) than tested TRs not passing ANOVA for any gene considered (n=464,220 unique TRs), which may reflect higher power to robustly detect nonlinear relationships for TRs that are more multi-allelic and therefore have a wider range of unique allele lengths available for testing. We additionally found that TRs passing ANOVA are enriched for homopolymers (Fisher’s Exact test two-sided *P*=1.10e-140, OR=1.71) and depleted for other repeat unit classes (**Supplementary Table 2**; **Methods**) compared to TRs not passing ANOVA for any gene tested, which may in part reflect the higher heterozygosity of these repeats resulting in greater power to detect nonlinear trends.

An example TR for which a nonlinear effect on gene expression was detected is a CCG_n_ repeat at hg38:chr3:19947266 associated with expression of the nearby gene *EFHB*. In the Geuvadis EUR cohort, this TR does not show a significant linear association with expression (two-sided *P*=0.61), but shows a significant quadratic effect (quadratic term two-sided *P*=3.25e-5) and a significantly better fit for the quadratic vs. linear model (ANOVA *P*=0.0013). Similar trends were observed in GTEx (linear *P*=0.99; quadratic *P*=1.1e-5; ANOVA *P*=2.0e-6). Visual inspection of the association between TR length and expression shows a bell-shaped trend with highest expression for alleles of 8-9 repeat copies and reduced expression at either longer or shorter alleles (**Fig. 4d**; **Supplementary Fig. 15**). Another example is a C_n_ repeat at hg38:chr10:70,473,982 associated with expression of *PALD1* (*KIAA1274*) for which a quadratic model is strongly favored in both Geuvadis (ANOVA *P*=8.4e-26) and GTEx (ANOVA *P*=0.0014). In this case, the median allele (copy number 10) is associated with lower expression compared to longer and shorter alleles (**Fig. 4e**; **Supplementary Fig. 16**).

Finally, we evaluated whether incorporating nonlinear effects of TRs could improve gene expression prediction models. In addition to the SNP-only and SNP+TR linear models above, we tested a third set of models for each gene containing SNPs as well as both linear and quadratic TR features for each TR included. As in our association testing models (**Methods**), the quadratic term is composed of the sum of squared allele lengths for the two alleles in each diploid individual. Overall, incorporation of quadratic TR effects resulted in improved adjusted R-squared values for 737 genes compared to the SNP-only and SNP+TR models. In most cases, improvements are modest, but 100 genes showed improvements of more than 0.01 (**Supplementary Table 1**). One of these is *PALD1*, for which a strong nonlinear effect of a C_n_ repeat was detected as described above, which showed an improvement in adjusted R-squared of 0.012 over the SNP+TR linear model and 0.069 over the SNP-only model. The LASSO model fit, which assigns a large coefficient to the quadratic term for this TR (0.11), closely matches the observed data and model fit from quadratic association tests (**Fig. 4e**). Taken together, our results show that although robust examples of nonlinear effects of TRs on expression exist, incorporation into expression prediction models, at least using available sample sizes, has minimal overall improvement on prediction performance for most genes (**Discussion**).

## Discussion

Here, we presented a framework for incorporating TRs into prediction models. Benchmarking our framework on simulated data demonstrated that including TRs can result in substantial improvement in predictive performance over SNP-only models, particularly at genes for which TRs with high effect sizes on expression are in weak to moderate LD with nearby SNPs. By applying this framework to gene expression for LCLs, we found that incorporating TRs results in simplified LASSO models with fewer non-zero coefficients, and provides especially large performance gains for a subset of 83 genes. For many genes, inclusion of TRs resulted in models that were more portable across ancestries, even when the SNP+TR model showed only a modest improvement in predictive performance over SNP-only models in the training European cohort. Finally, we identified tens of thousands of examples of nonlinear associations between TR copy number and expression, with 252 associations that could only be detected when applying nonlinear models. We highlight several strong examples of nonlinear effects, including a C_n_ repeat associated with *PALD1*, and show that incorporating the nonlinear effect resulted in an improved gene expression prediction model.

We hypothesize that the gains in performance, portability, and simplification of prediction models is due to the direct inclusion of additional causal variants as predictors. Current polygenic scores including gene expression prediction models primarily rely on common SNPs. In many cases, the variants included are not causal, but rather serve as LD proxies for other underlying causal variants. As a result, they may achieve low performance in cases where causal variants are not well tagged, and performance degrades when predicting on individuals of other ancestries with different LD structures. Recent studies have demonstrated a widespread impact of TR variation on gene expression variability^10,28^. In many cases, these TRs are highly polymorphic and in weak to modest LD with individual nearby variants, although can often be imputed to high accuracy using combinations of SNPs^20^. As a result, LASSO prediction models may need to include multiple SNPs to achieve the same performance that can be achieved by including a single TR. Further, if a TR included in the model is truly causal, its predictive power is likely to remain high even when applying models to ancestries with different LD structures, as many TR effects on expression have been observed to be shared across ancestries^25^.

Our results have important implications for the design of polygenic risk scores (PRSs), which assess an individual’s disease risk or other complex trait values based on their genetic variants. Similar to gene expression prediction models, standard frameworks^31–33^ typically rely on common SNPs, and have shown low portability across ancestries with different LD structures and allele frequencies^34^. Beyond gene expression, recent studies have implicated highly polymorphic TRs as a major source of high impact causal variants across a range of medically relevant complex traits^7,17–19^. Overall, this suggests incorporating TRs into PRS will have similar gains to those observed here for gene expression prediction models.

Our study faced several limitations. First, the Geuvadis dataset only had a sufficiently large sample size to train across individuals of European ancestry. While inclusion of TRs resulted in improved portability for some genes, TRs often show population-specific allele ranges and frequencies, whose effects will not be captured by our models here. Expanding to additional datasets such as the multi-ethnic study of atherosclerosis (MESA) dataset which contains 574 European samples and 224 African samples of monocyte tissue^35^ may result in improved models for non-European samples in future studies. Third, we only considered a single cell type (LCLs). Still, we anticipate the overall trends observed here will generalize to other settings, and that expanding to more cell types and tissues will identify additional genes for which incorporating TRs provides a major improvement in performance of gene expression prediction models.

Finally, our models are based on a limited sample size (n=359 samples from Geuvadis). This limits our ability to accurately model variants with low effect sizes and contributes to high variance in expression predictions. We also anticipate that low sample size reduces our ability to detect and model nonlinear effects. Despite evidence from our recent work on massively parallel reporter assays (MPRA)^28^ that nonlinear effects of TRs on expression may be widespread, we found only modest improvement in model performance when including nonlinear TR features. One potential explanation is that our use of a LASSO model with quadratic TR length effects is insufficient to capture some types of nonlinear trends. Other model classes such as random forests, gradient boosting, or neural networks may better capture more complex relationships between TR length and expression. However, these models are more expensive to train and require larger sample sizes to avoid overfitting. Another challenge in modeling nonlinear effects at low sample sizes is that we may not observe a sufficient number of different TR lengths to reliably infer nonlinear trends. For example, although we observed nonlinear trends when considering more than a dozen different repeat lengths at particular locus in an MPRA context^28^, if only two or three alleles are observed they may still be best fit by a linear model.

Overall, our results demonstrate that incorporating TRs into polygenic prediction models can result in improved and simpler models with improved portability across ancestries. These results suggest that including additional potential sources of causal genetic variation beyond common SNPs in PRS and related methods is likely to result in improved prediction models for a range of traits.

## Methods

### Geuvadis dataset preprocessing

Gene expression for EBV-transformed lymphocytes was obtained from the Geuvadis project for European (super population code EUR, n = 359) and YRI (super population code AFR, n = 89) individuals^23^ (**URLs**). We used gene annotations based on Gencode v12. Gene coordinates were lifted over from hg19 to hg38 using the UCSC liftOver tool^36^. We restricted our analyses to protein coding genes. We further removed genes in segmental duplications (based on UCSC genomicSuperDups table), genes with expression below 0.1 RPKM in more than 10 samples, and genes for which liftOver failed or resulted in coordinates on an ALT contig of hg38. After these filters, 12,605 protein-coding genes remained for analysis. Expression values for each gene were first quantile normalized across samples to ensure all samples follow similar distributions, then values for each gene were inverse rank normalized to follow a standard normal distribution.

SNP genotypes for these individuals based on high coverage whole genome sequencing of 1000 Genomes Project samples^22^ and sex information were obtained from the IGSR data portal (**URLs**). We filtered all SNPs using plink2 version v2.0.0-a.6.9LM with a minor allele frequency less than 0.05 and failing Hardy-Weinberg Equilibrium (*P*<0.001). Additionally, we removed duplicate SNP entries (those with the same chromosome and position) keeping the first SNP listed in the given position. Genotype principal components (PCs) were computed from 8,975,030 autosomal SNPs using scikit-learn 1.7.2 separately for European and Yoruban individuals present in Geuvadis. Prior to downstream analyses we regressed out the top 5 genotype principal components (PCs), sex, and PEER factors^37^. PEER factors were computed using normalized expression values. We included 36 PEER factors for Europeans and 9 for Yorubans based on our previous finding that sample_size/10 PEER factors optimized discovery of TR-gene expression associations^10^. TR genotypes for 1.7 million loci were obtained using EnsembleTR as described previously^25^ (**URLs**). We filtered TR loci with two or fewer unique alleles and with a heterozygosity less than 0.10.

### Simulation analysis

To evaluate the gene expression model framework described below, we generated simulated gene expression values to assess whether incorporating TRs would improve model prediction accuracy. In each case, we first identified causal SNPs and/or TRs within a 500kb window around a given gene and assigned each causal variant (SNP or TR) an effect size. Simulations are based on real genotype data for the 359 European samples in the Geuvadis cohort to ensure they recapitulate realistic LD patterns. The genotypes were Z-normalized before being passed to use for gene expression simulation and prediction. In order to calculate LD in the below analyses we used the Pearson product-moment correlation coefficients from numpy version 2.2.6.

We simulated two different scenarios. In the first, we used a polygenic architecture in which *cis*-effects were spread equally across up to 4 SNPs and 4 TRs (using genotypes corresponding to the window around gene *HEXDC*). Unless otherwise specified, causal variants were chosen at random every simulation round to determine how models would behave on average and capture variants across different causal variant sets. We additionally performed simulations (**Supplementary Fig. 2-3**) in which causal TRs were chosen specifically to have either low or high LD with *cis*-SNPs. Across all rounds for this first scenario, causal effect sizes were chosen such that the sum of squared effect sizes is 0.8, corresponding to a *cis*-heritability of approximately 80% (assuming no LD between causal variants).

In the second, we simulated a single causal TR (using genotypes corresponding to the window around gene *TIMM10*). For this we first chose the AT_n_ TR at hg38:chr11:57,528,484 as the causal variant, based on our previous observation that this TR is a strong eTR^25^. We varied the squared TR effect size from 0 to 0.8. We performed additional sets of simulations using other TRs in this region as the single causal TR to assess the impact of LD with the best tag SNP on performance gain for SNP+TR vs. SNP only models (**Supplementary Fig. 4**).

For each scenario we simulated expression values using the equation: 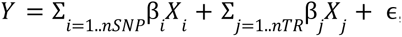 where *Y* is a vector of simulated values, β is the effect size of SNP *i*, *X_i_* is the normalized SNP genotypes for SNP *i*, *nSNP* is the number of simulated causal SNPs, β*_j_* is the effect size of TR *j*, *X_j_* is the normalized genotypes for TR *j*, *nTR* is the number of simulated causal TRs, and ɛ is the error term drawn from *N*(0, σ*_e_*^2^) for each sample, where 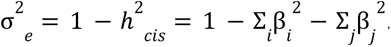. TR genotypes are encoded using the total number of copies of whole motifs in tandem. Simulated expression values were standardized to have mean 0 and variance 1 prior to downstream analysis. We repeated each simulation setting 100 times. Model training and evaluation of SNP and SNP+TR models for each simulated dataset were computed using the same procedure as described for real data below.

### Computing gene expression models

We trained models to predict expression for each gene based on *cis*-variants using LASSO regression implemented in the scikit-learn package version 1.7.2 (**ULRs**) with non-default parameter max_iterations=50000 and tolerance 1e-4. For each gene, we considered SNPs and TRs from an interval encompassing the gene boundaries +/- 500kb. Prior to training genotypes each variant was standardized to have mean zero and variance 1. We trained two separate models for each gene, including either SNPs only or SNPs+TRs. For each model, the alpha parameter for LASSO was tuned using the GridSearchCV class from scikit-learn to train models using 3-fold cross validation with n_repeats=2. Tuning was performed in two steps, the first performed a broad search using a range of 0.1-0.9 with step 0.04. The second performed a higher resolution search using the value learned in the first round +/- 0.09, with minimum allowable alpha 0.001, searching 51 possible values. Due to our relatively low sample size (n=359 European samples), we repeated the training process above 20 times to assess variability in performance across runs. For each gene we summarized the average R-squared, adjusted R-squared, and standard deviation across the 20 iterations. We tested whether SNP+TR models resulted in significant gains in adjusted R-squared over SNP-only models using 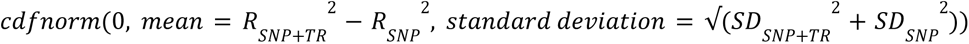, where 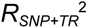 and 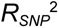 give the mean adjusted R-squared for the 20 SNP+TR and SNP-only models, respectively, and the SD values give the standard deviation across all models.

Heritability values were obtained using GCTA and estimated using Hapmap SNPs as described in Akamatsu *et al.*^38^. Because heritability estimation is performed only based on SNPs, we anticipate heritability values are in some cases underestimates of the true value due to *cis*-variation.

### Validating gene expression prediction models in GTEx

We utilized data from the GTEx Project^30^ v8 to validate gene expression prediction models. SNP genotypes were obtained through dbGaP accession number phs000424.v8.p2. TR genotypes were imputed from SNPs via Beagle 5.4^39^ using the EnsembleTR reference panel^25^. Expression data for EBV transformed lymphocytes was obtained from GTEx_Analysis_2017-06-05_v8_RNASeQCv1.1.9_gene_tpm.gct on the GTEx portal. We filtered genes that had a TPM < 1 in 90% or more of the samples and then regressed out the top 5 PCs and 11 PEER factors. Of the 147 samples that had expression data for EBV transformed lymphocytes, we filtered samples that had any missing genotypes for any SNPs receiving non-zero coefficients in any gene expression prediction models across all genes resulting in 131 total samples. We used PCA on SNP genotypes found from GTEx_Analysis_v8_eQTL_covariates.tar.gz on the GTEx portal and 1000 Genomes to obtain genetically predicted ancestry for each individual. In particular, we identified samples with PC1 > −0.014 and PC2 > 0.01 as EUR, with remaining samples with available expression predicted as AFR (**Supplementary Fig. 9**). This resulted in 109 EUR and 22 AFR samples.

We used genotyped SNPs and imputed TRs for these samples, standardized to have mean 0 and variance 1 as in the training data, to obtain predicted gene expression values based on models trained on EUR samples in Geuvadis. Because our TR imputation panel contains a subset of 1M TRs than the total 1.7M genotyped TR calls for Geuvadis, not all TRs included in these models were available in GTEx. Of the 1951 gene models with non-zero TR coefficients, 1,588 had genotypes for at least one relevant TR in GTEx. Of these, 53 genes were not present in the GTEx EBV transformed lymphocyte expression data, leaving 1,530 genes for validation. Among these remaining genes, not all TRs with non-zero coefficients had imputed genotypes available. We therefore stratified results on the total weight of TRs included, computed as the sum of the absolute value of coefficients for TRs with genotypes available in the GTEx cohort (**Fig. 2e**).

### Cross population gene expression models

We used gene expression prediction models trained in the EUR cohort to predict expression for individuals from the YRI cohort of Geuvadis based on SNP and TR genotypes. As in the EUR analysis, genotypes were Z-normalized to have mean 0 variance 1. We compared predicted expression values to observed expression values for each sample, adjusted for 9 PEER factors, sex, and the top 5 genotype PCs computed from YRI samples. Prior to comparing to predicted values, adjusted expression values were normalized to have mean 0 and variance 1. For each gene, we computed the Pearson *r^2^* between predicted and observed expression values.

### Linear and nonlinear association testing

We performed linear and nonlinear association testing for TRs against nearby genes within the Geuvadis and GTEx datasets described above. In Geuvadis, we utilized the same covariates as described above and in GTEx we used sex, the top 5 PCs, and 11 PEER factors as covariates. For linear models, for each TR-gene pair, considering TRs within 500kb of the boundaries of each gene, we fit models of the form *Y* = β(*x*_1_ + *x*_2_ ) + γ*C* + ɛ, where *x*_1_ + *x*_2_ gives the sum of the TR lengths (Z-normalized to have mean 0 and variance 1) of the two alleles of each individual, and other variables are defined as above. For nonlinear association testing, we fit the model 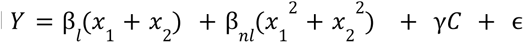 (in this case TR lengths are not normalized). This model assumes additive contributions to expression from the two chromosome copies, with each following a model 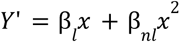, where *Y*’ is the expression coming from a single chromosome copy and *x* is the number of repeats on that same chromosome copy. We computed P-values for each regression coefficient (β for linear models, β*_l_* and β*_nl_* for nonlinear models) using the OLS function from statsmodels version 0.14.6. We additionally performed an ANOVA test using statsmodels anova_lm python function to determine whether nonlinear models provide an improved fit for each gene. To identify genome-wide significant signals for either the linear or quadratic, we first identified the TR with the top P-value for each gene. These were adjusted for the number of tests using a Bonferroni correction. Adjusted top P-values were then used as input to a Benjamini-Hochberg FDR correction to identify associations that are significant at a gene-level FDR of 5%. A similar procedure was applied to ANOVA p-values comparing linear vs. nonlinear models.

To incorporate nonlinear effects into gene expression models, we similarly included an additional term *X*^2^_1_+ *x*_2_^2^ for each TR. However, unlike in our nonlinear association testing above, we normalized the sum of the squares of the TR alleles to have mean 0 variance 1. Normalization of these values is necessary in order to ensure the regularization term learned throughout the LASSO training process is generalizable to all features.

### Nonlinear association enrichment testing

We tested whether TRs showing significant nonlinear associations with expression were enriched for certain properties compared to all tested TRs. For each TR, we identified the gene for which it had the best corrected ANOVA p-value. For each repeat unit length and/or motif sequence, we performed a separate Fisher exact test using TRs with best adjusted ANOVA p-values below 0.05 as the foreground and all other TRs as the background. We only performed tests for categories (repeat unit length or sequence) for which the foreground contained at least 10 TRs with that annotation. Fisher exact tests were performed in Python using scipy version 1.15.2.

## Data availability

Geuvadis datasets and 1000GP genotypes used in this study are available from the 1000GP website [https://www.internationalgenome.org/data-portal/data-collection/geuvadis] and [https://www.internationalgenome.org/data-portal/data-collection/30x-grch38]. Analyses are based on the GRCh38 reference genome [https://storage.googleapis.com/genomics-public-data/resources/broad/hg38/v0/Homo_sapiens_assembly38.fa <u>sta</u>]. GTEx data was obtained from dbGaP accession number phs000424.v8.p2 and from the GTEx portal website [https://www.gtexportal.org/home/downloads/adult-gtex].

## Code availability

Main analysis scripts and workflows are available on Github: https://github.com/mlamkin7/TRGeneExprModel_workflows

## Data Availability

Geuvadis datasets and 1000GP genotypes used in this study are available from the 1000GP website [https://www.internationalgenome.org/data-portal/data-collection/geuvadis] and [https://www.internationalgenome.org/data-portal/data-collection/30x-grch38]. Analyses are based on the GRCh38 reference genome [https://storage.googleapis.com/genomics-public-data/resources/broad/hg38/v0/Homo_sapiens_assembly38.fasta]. GTEx data was obtained from dbGaP accession number phs000424.v8.p2 and from the GTEx portal website [https://www.gtexportal.org/home/downloads/adult-gtex].

## Acknowledgments

This work was partially supported by NIH/NHGRI grants R01HG010885 and RM1HG011558 (M.G.), National Science Foundation (NSF) Award Number 2336469 (T.A.) and by NIH/NHGRI grant R01HG013671 (T.A.).

## URLs

Geuvadis expression data: https://www.internationalgenome.org/data-portal/data-collection/geuvadis EnsembleTR genotype calls: https://github.com/gymrek-lab/EnsembleTR

High-coverage WGS of 1000 Genomes samples: https://www.internationalgenome.org/data-portal/data-collection/30x-grch38

1000 Genomes Project sample metadata: https://www.internationalgenome.org/data-portal/sample

## Author contributions

M.L. performed analyses and wrote the manuscript. Y.L. performed preprocessing of Geuvadis and GTEx datasets. K.A. computed *cis*-heritability values for each gene. T.A. helped supervise analyses. M.G. conceived the study, supervised analyses, helped write the manuscript, and obtained funding.

