## Supplementary Figures for "Incorporating tandem repeats improves gene expression prediction models"

Supplementary Figure 1

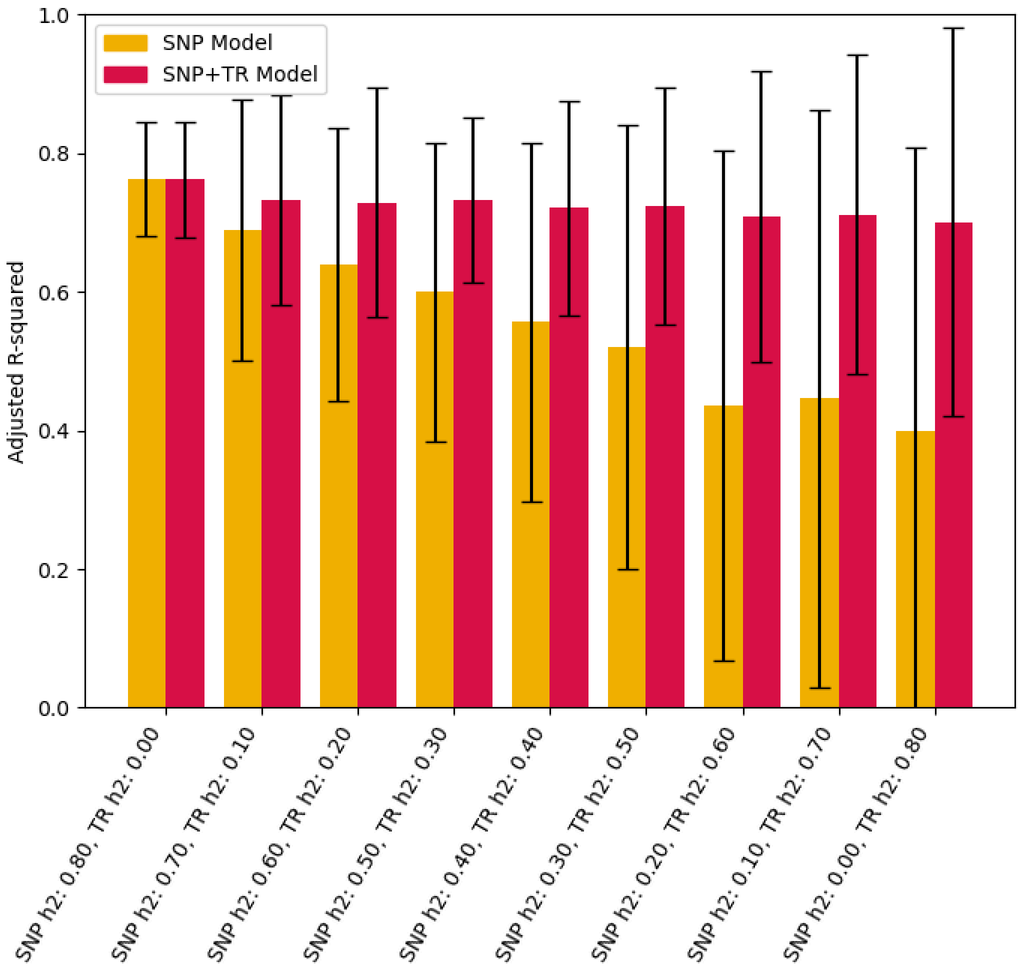


**Evaluation of gene expression models on simulated expression based on 2 causal SNPs and 2 causal TRs.** Simulations are the same as in **Fig. 1b**, but with two causal SNPs and TRs rather than 4, with a total *cis* heritability of 0.8. Bars show performance of the SNP only (gold) and SNP+TR (red) gene expression prediction models measured using adjusted R-squared based on 3-fold cross validation. The *cis* heritability driven by SNPs vs. TRs is annotated for each setting on the x-axis. Error bars show 95% confidence intervals across 100 simulation rounds.

Supplementary Figure 2

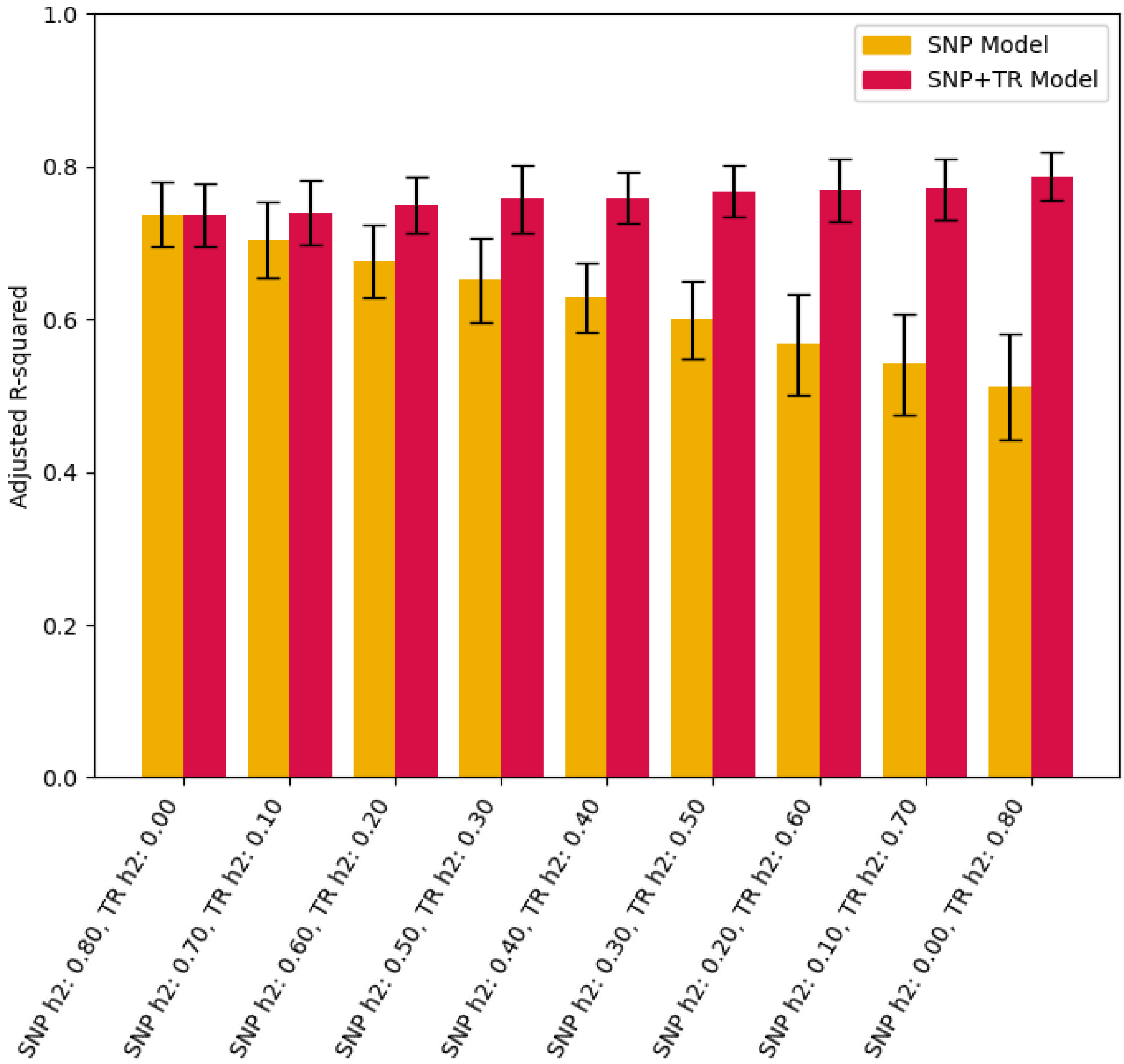


**Evaluation of gene expression models on simulated expression based on multiple causal SNPs and TRs for which TRs are in low LD with SNPs.** Simulations and the plot are the same as in **Fig. 1b**, except TRs were chosen such that they have low LD with any *cis* SNP. Specifically, variants include: (SNP: hg38:chr17:82,044,620, TR: hg38:chr17:82,071,841, LD: 0.0589), (SNP: hg38:chr17:82,504,059, TR: hg38:chr17:82,574,322, LD: 0.0340), (SNP: hg38:chr17:82,412,549, TR: hg38:chr17:82,422,496, LD: 0.0991), and (SNP: hg38:chr17:82,503,792, TR: hg38:chr17:82,123,123, LD: 0.0283). LD values give $r^2$ for the best tag SNP for each TR.

#### Supplementary Figure 3

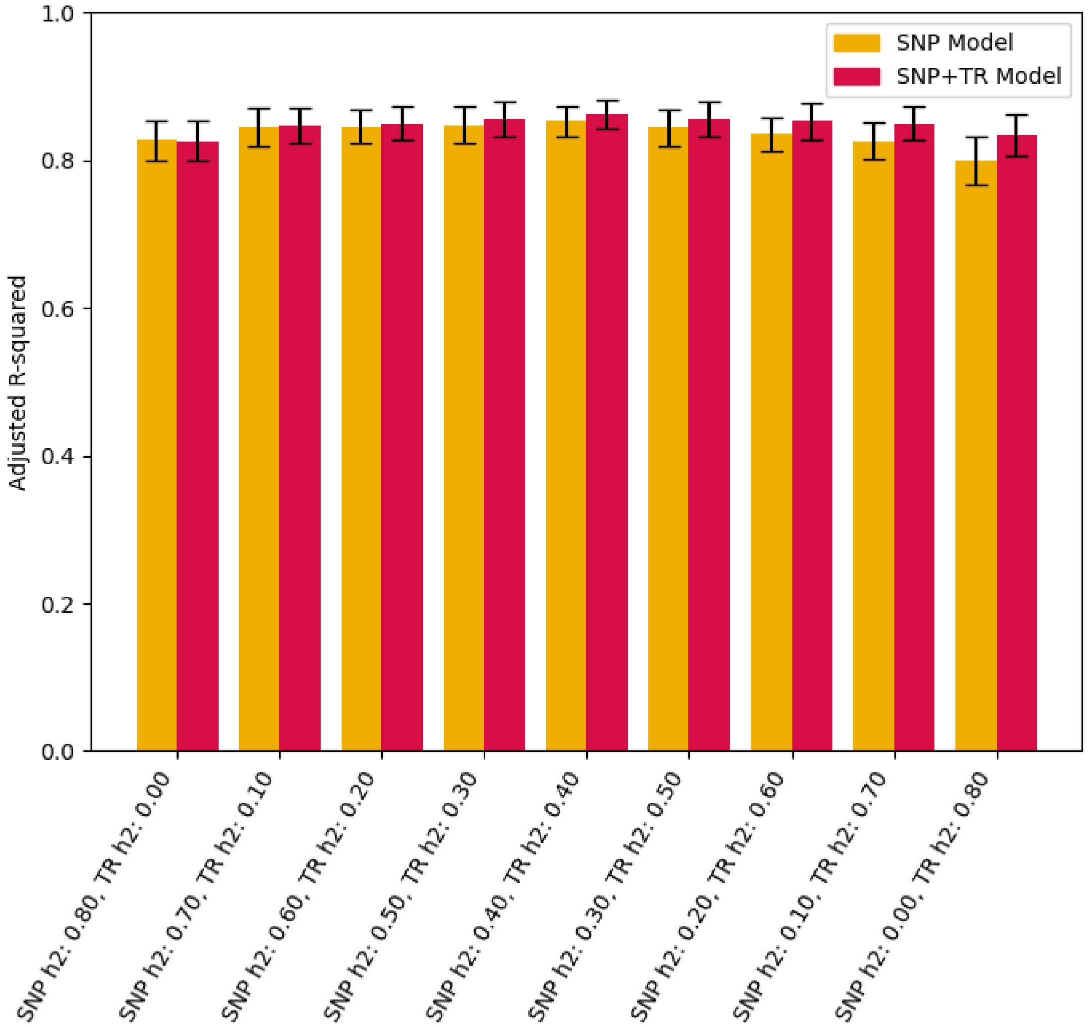


**Evaluation of gene expression models on simulated expression based on multiple causal SNPs and TRs for which TRs are in high LD with SNPs.** Simulations and the plot are the same as in **Fig. 1b**, except TRs were chosen such that they have high LD with any *cis* SNP. Specifically, variants include: (SNP: hg38:chr17:82,596,912, TR: hg38:chr17:82,603,720, LD: 0.9282), (SNP: hg38:chr17:82,872,389, TR: hg38:chr17:82,879,060, LD: 0.9409), (SNP: hg38:chr17:82,101,611, TR: hg38:chr17:82,210,518, LD: 0.9122), and (SNP: hg38:chr17:82,919,244, TR: hg38:chr17:82,920,537, LD: 0.9072). LD values give $r^2$ for the best tag SNP for each TR.

Supplementary Figure 4

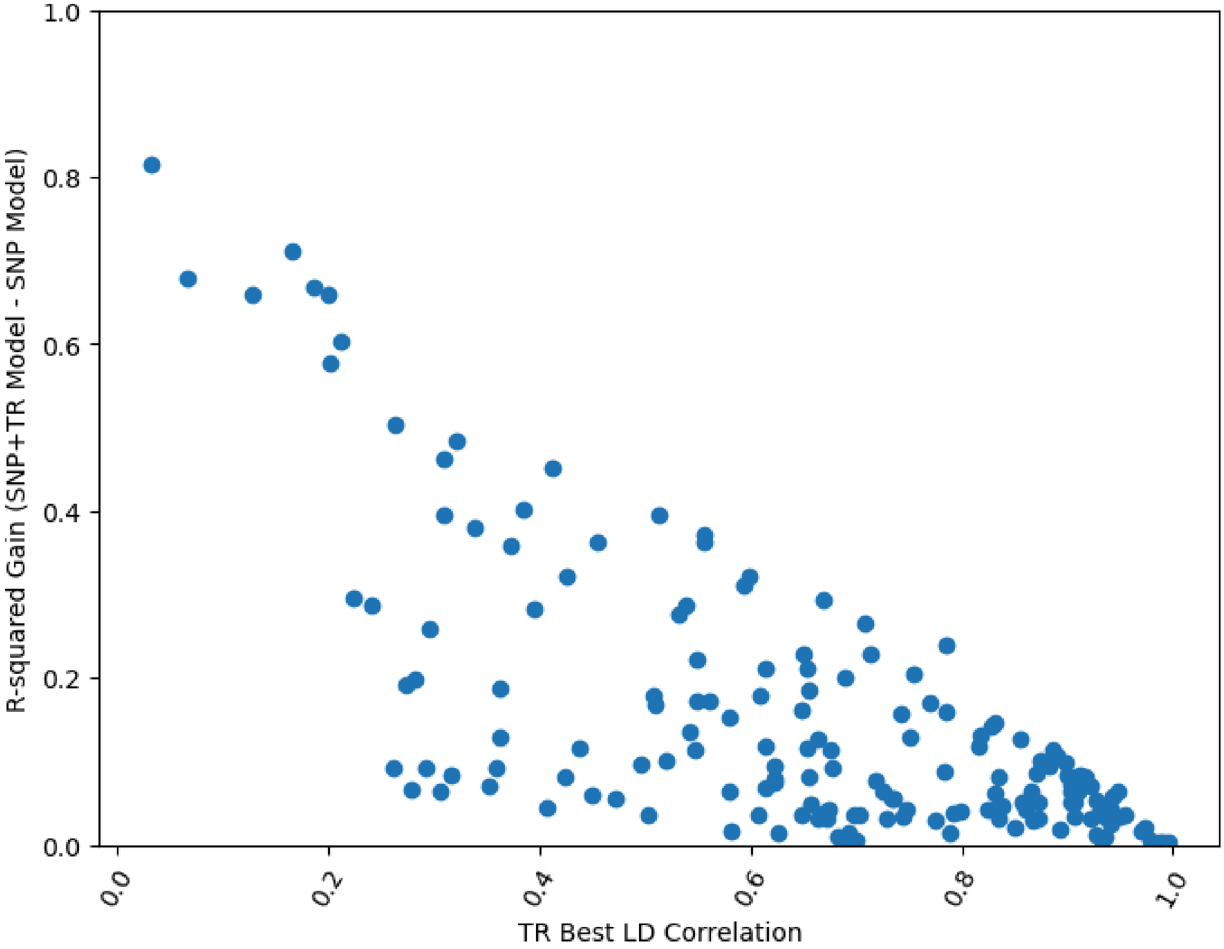


**Impact of SNP-TR LD correlation on simulated gain.** We repeated simulations with a single causal TR for *TIMM10* for all 180 TRs within 500kb of the gene boundaries, in each case treating one TR as a single causal variant explaining 80% of *cis* heritability of expression. The x-axis gives the maximum pairwise LD ($r^2$) between the causal TR and any of the 931 common SNPs in the region. The y-axis shows the gain in adjusted R-squared for SNP+TR models vs. SNP only models for each simulation.

#### Supplementary Figure 5

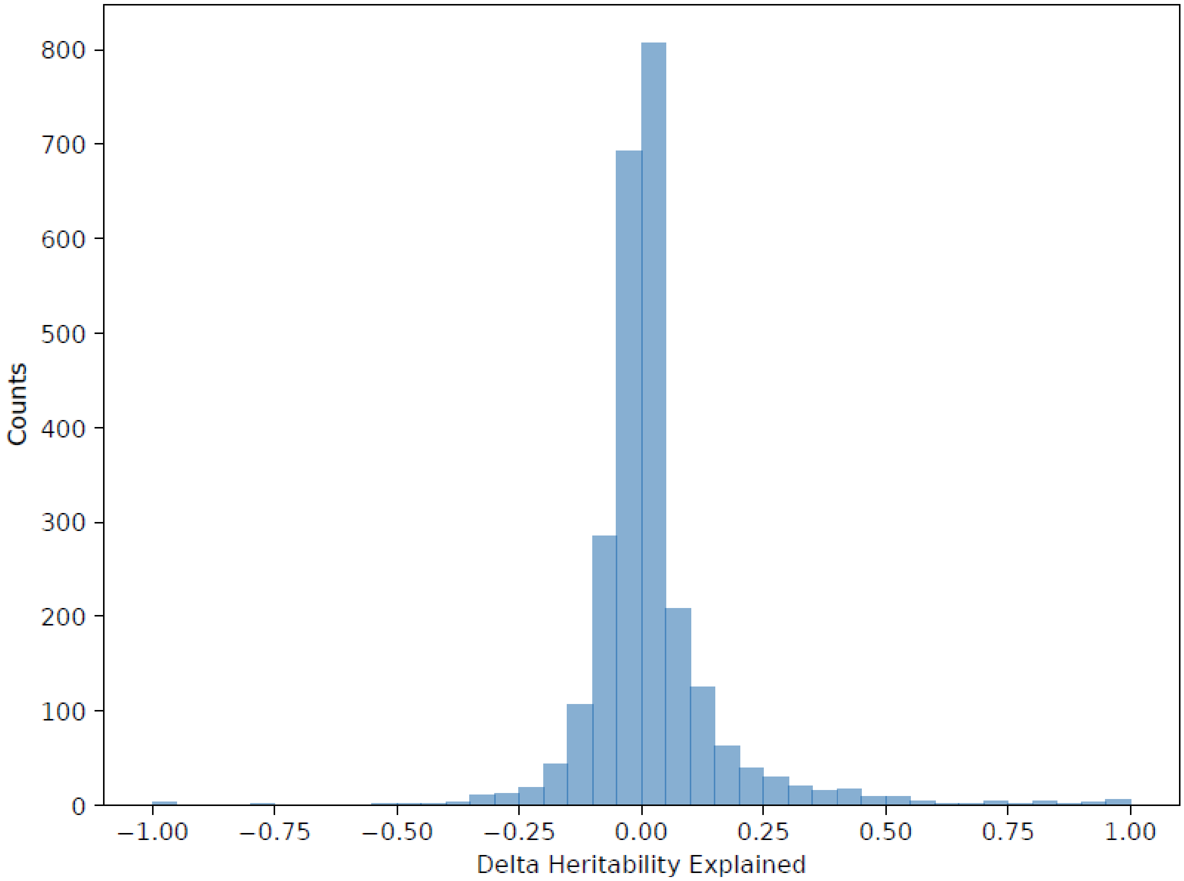


**Distribution of the difference in *cis* heritability explained using SNP+TR vs. SNP-only gene expression prediction models.** Heritability explained is defined as the adjusted R-squared of the model divided by the total estimated *cis* heritability for each gene (**Methods**). Data is based on n=2,553 genes for which LASSO models were successfully trained.

### Supplementary Figure 6

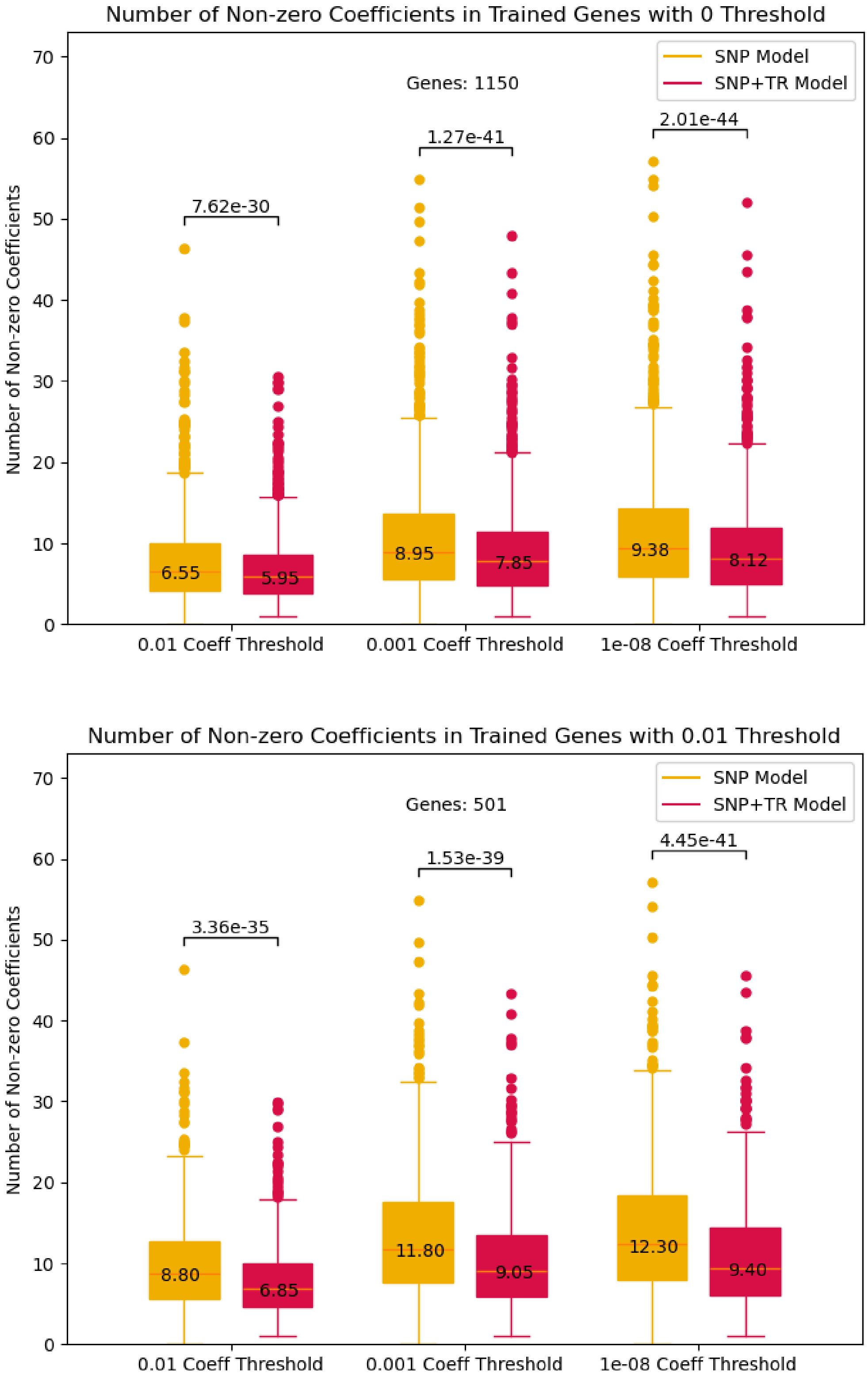

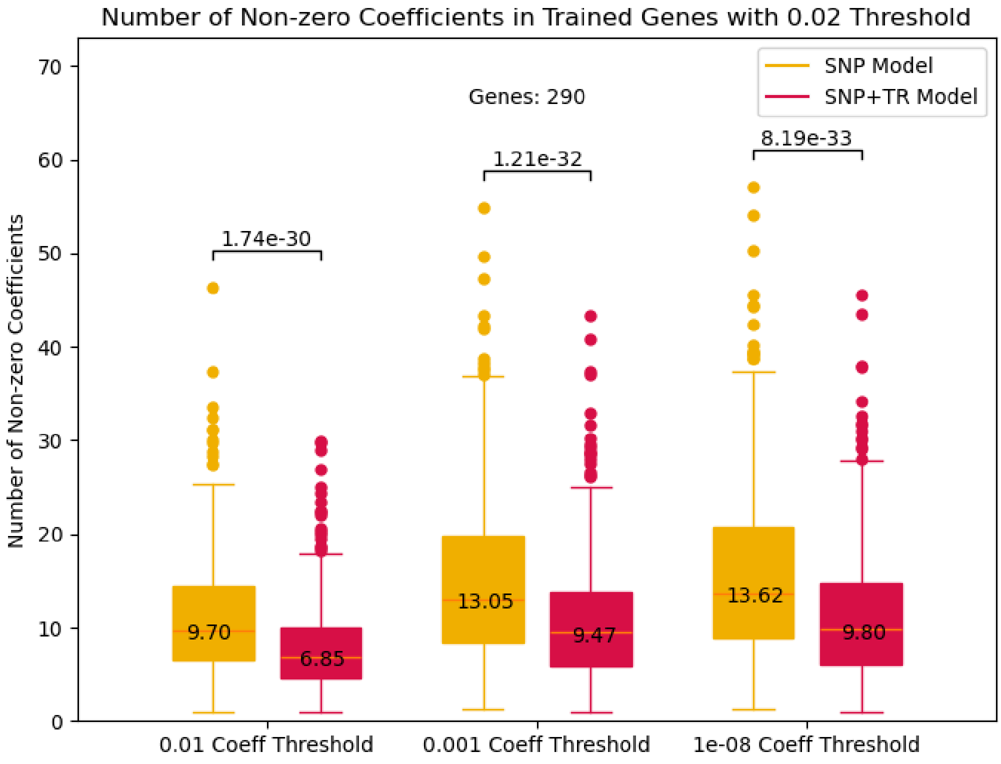

Number of Non-zero Coefficients in Trained Genes with 0.02 Threshold
SNP Model
SNP+TR Model
Genes: 290
1.74e-30
1.21e-32
8.19e-33
9.70
6.85
13.05
9.47
13.62
9.80
Number of Non-zero Coefficients
0.01 Coeff Threshold
0.001 Coeff Threshold
1e-08 Coeff Threshold


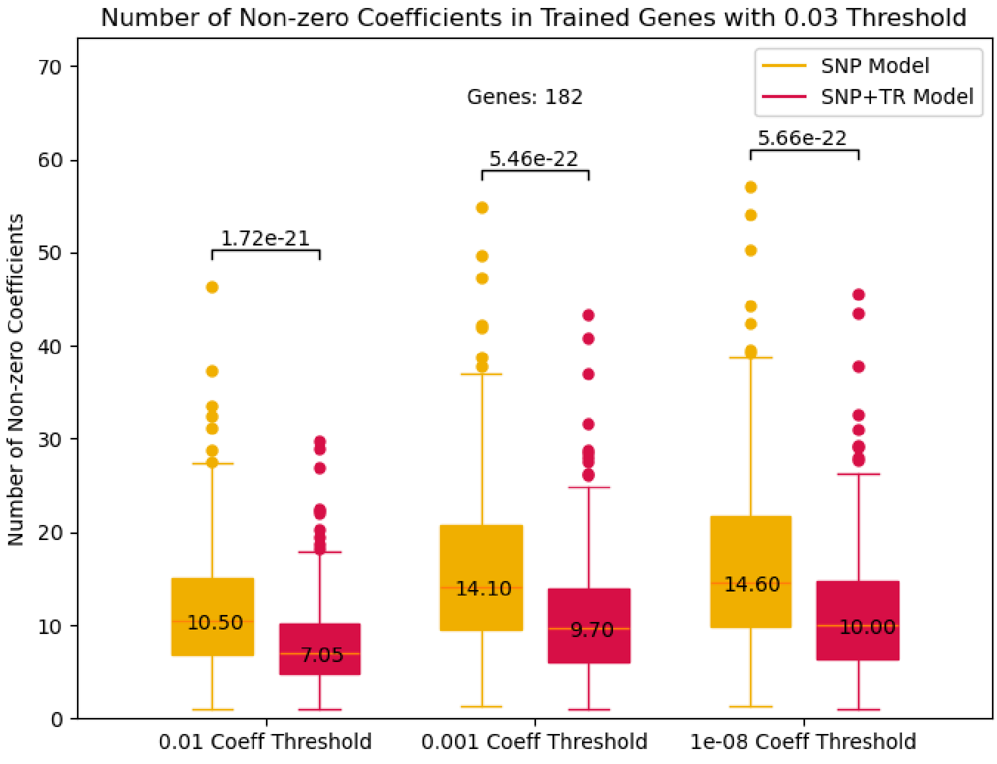

Number of Non-zero Coefficients in Trained Genes with 0.03 Threshold
SNP Model
SNP+TR Model
Genes: 182
1.72e-21
5.46e-22
5.66e-22
10.50
7.05
14.10
9.70
14.60
10.00
Number of Non-zero Coefficients
0.01 Coeff Threshold
0.001 Coeff Threshold
1e-08 Coeff Threshold

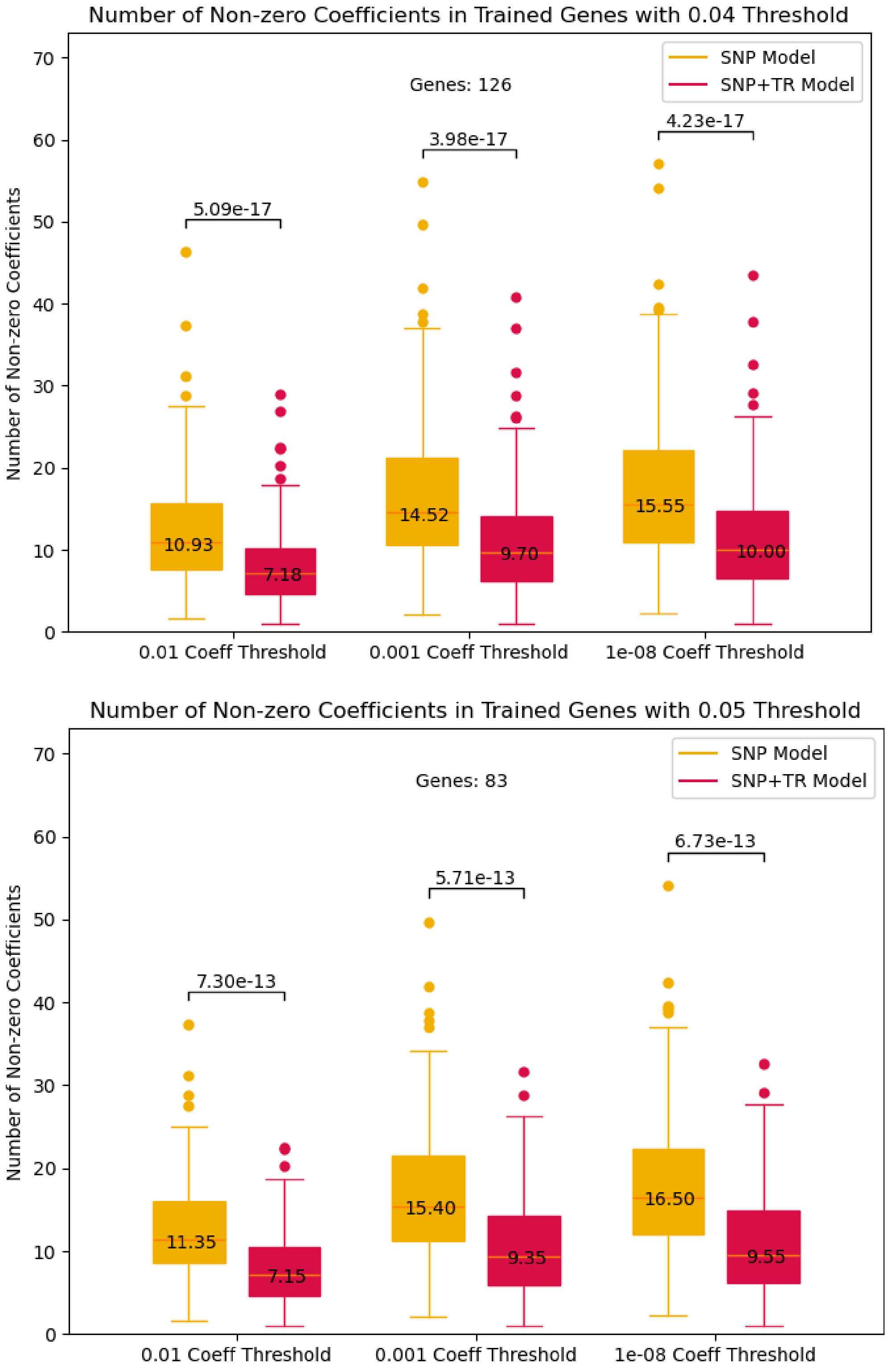


**Distribution of the number of non-zero coefficients in the SNP-only vs. SNP+TR gene expression prediction models.** Gold=SNP-only models; red=SNP+TR models. Each panel is restricted to genes for which SNP+TR models showed a gain in adjusted R-squared above the threshold listed in the title of each plot (either 0, 0.01, 0.02, 0.03, 0.04, or 0.05). Within each panel, x-axis values give the LASSO coefficient value ($\epsilon$) above which a variant was considered to have non-zero weight.The number of genes considered in each plot (ranging from 1,150 to 83) is annotated. For each setting, the one-sided P-value from a Wilcoxon signed-rank test comparing the two distributions is annotated. For each boxplot, horizontal lines show median values, boxes span from the 25th percentile (Q1) to the 75th percentile (Q3). Whiskers extend to $Q1 - 1.5 \times IQR$ (bottom) and $Q3 + 1.5 \times IQR$ (top), where IQR is the interquartile range (Q3–Q1).

Supplementary Figure 7

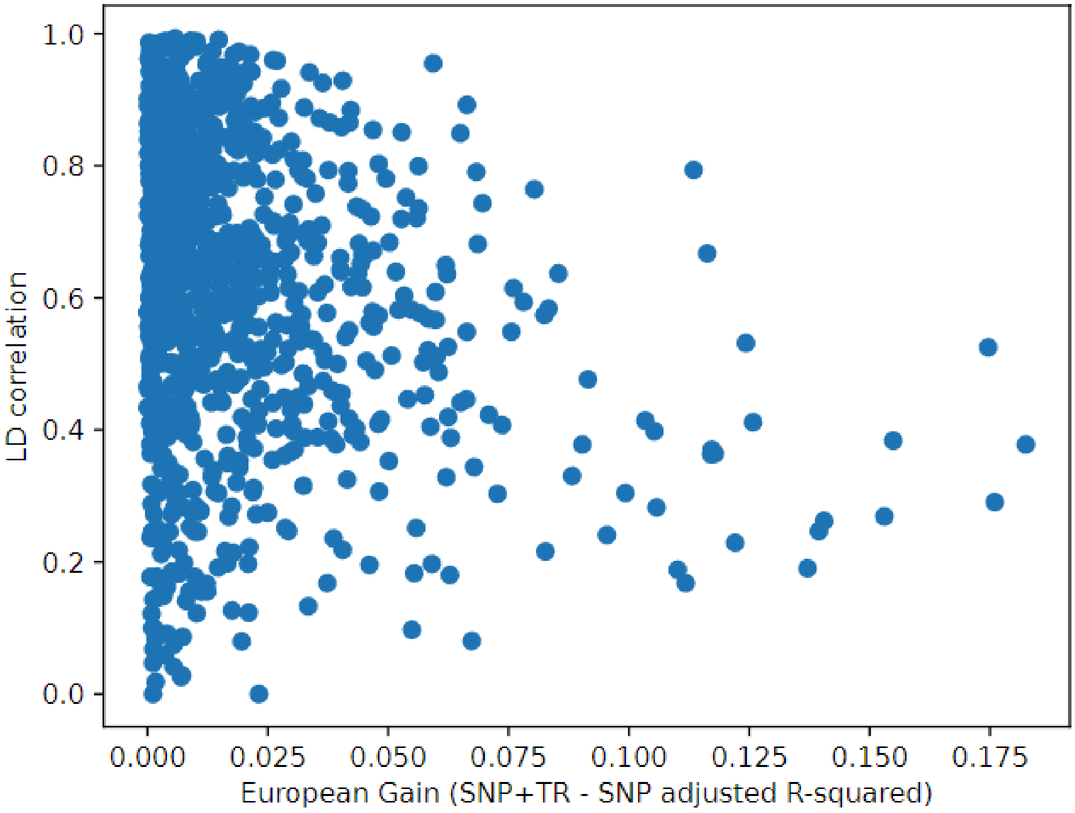


**Correlation of performance gain of SNP+TR models with SNP-TR LD.** The x-axis gives the difference in adjusted R-squared for SNP+TR vs. SNP-only models for each gene for which adding TRs improved the model and at least one TR had a non-zero model coefficient (n=1,009 genes). The y-axis gives the LD ($r^2$) of the TR with the highest absolute value coefficient and its best tag SNP considered in the model.

### Supplementary Figure 8

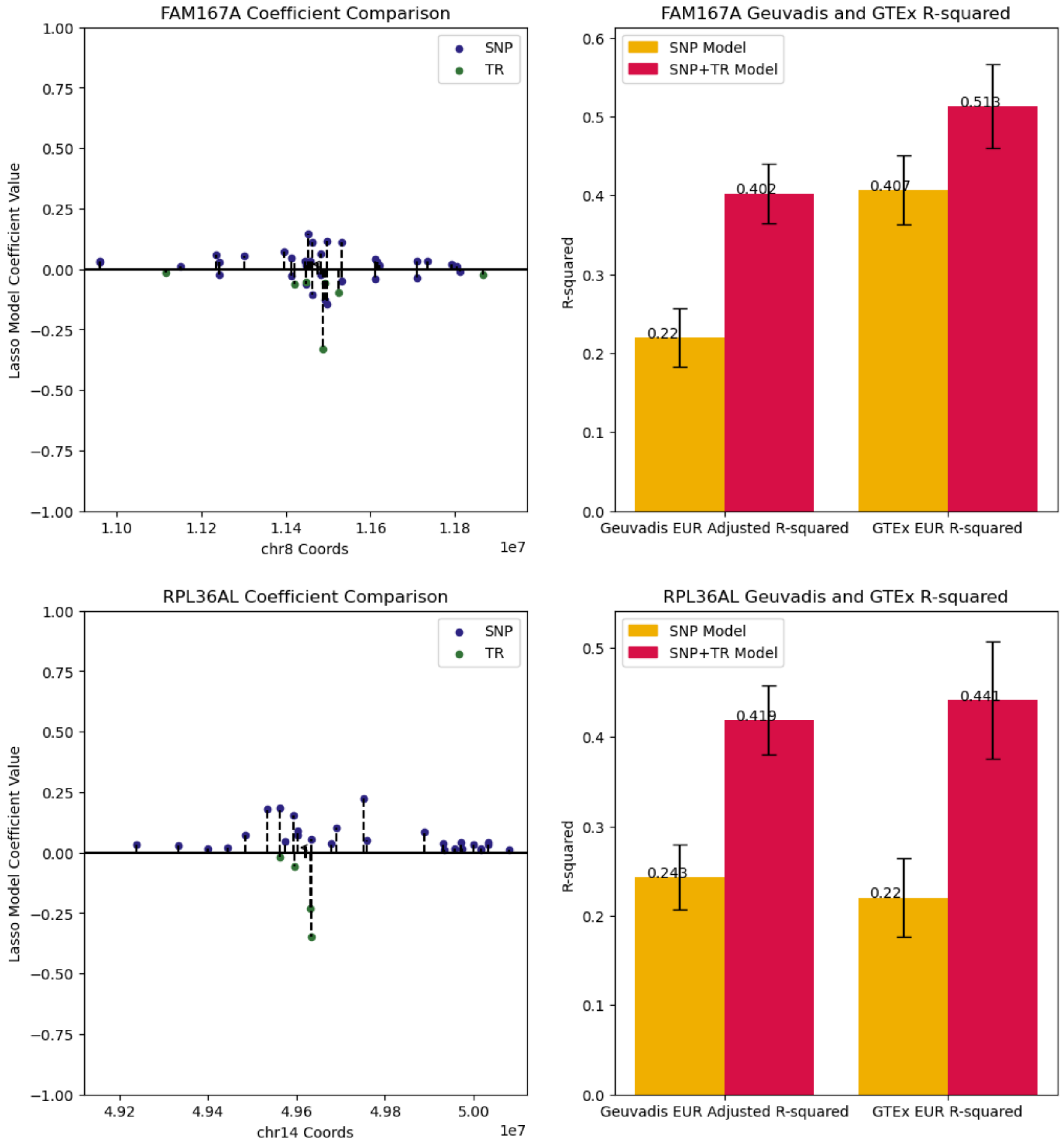

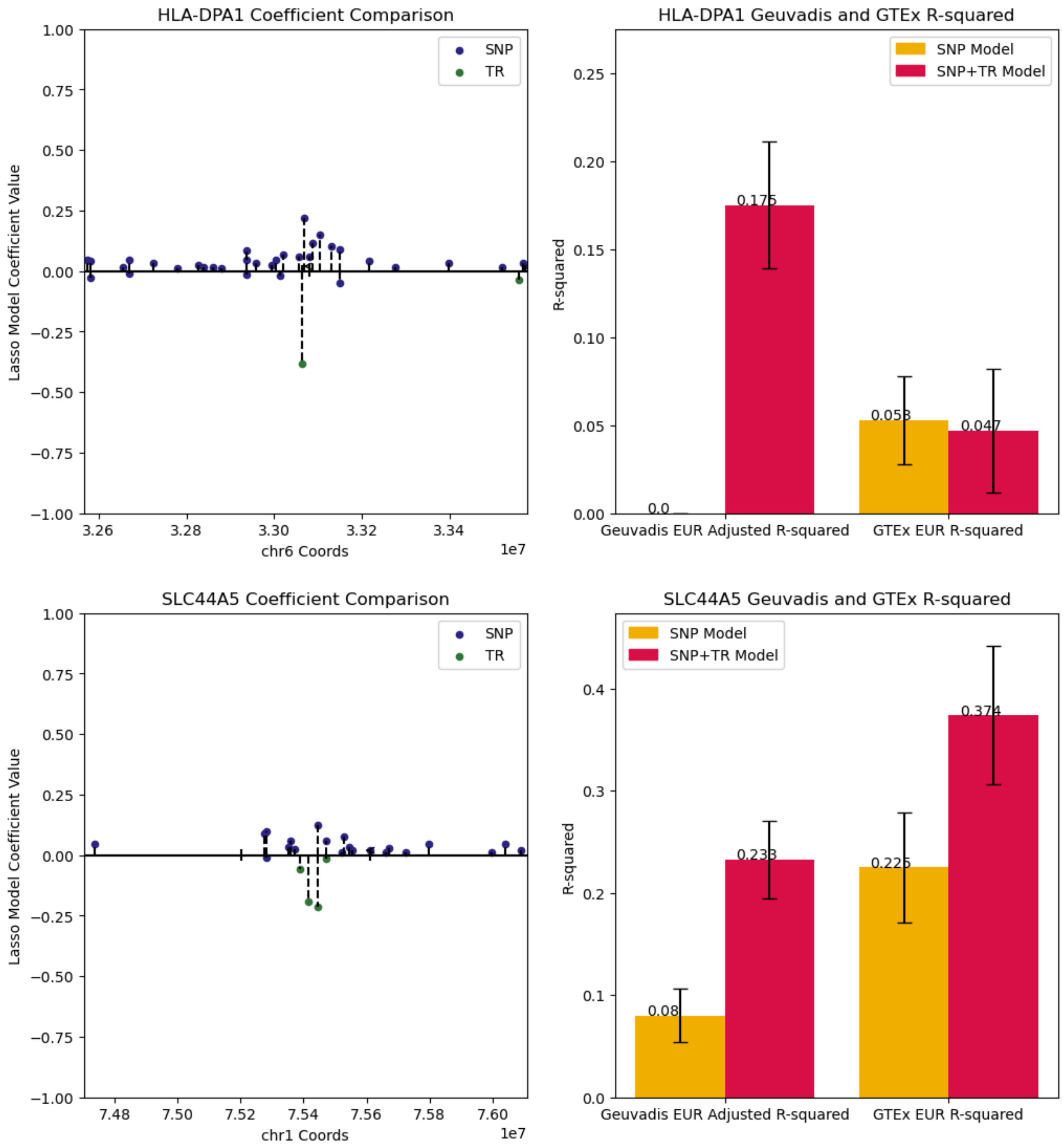

HLA-DPA1 Coefficient Comparison
Lasso Model Coefficient Value
chr6 Coords
1e7
SNP
TR
HLA-DPA1 Geuvadis and GTEx R-squared
R-squared
SNP Model
SNP+TR Model
0.0
0.175
0.053
0.047
Geuvadis EUR Adjusted R-squared
GTEx EUR R-squared
SLC44A5 Coefficient Comparison
Lasso Model Coefficient Value
chr1 Coords
1e7
SNP
TR
SLC44A5 Geuvadis and GTEx R-squared
R-squared
SNP Model
SNP+TR Model
0.08
0.233
0.225
0.374
Geuvadis EUR Adjusted R-squared
GTEx EUR R-squared

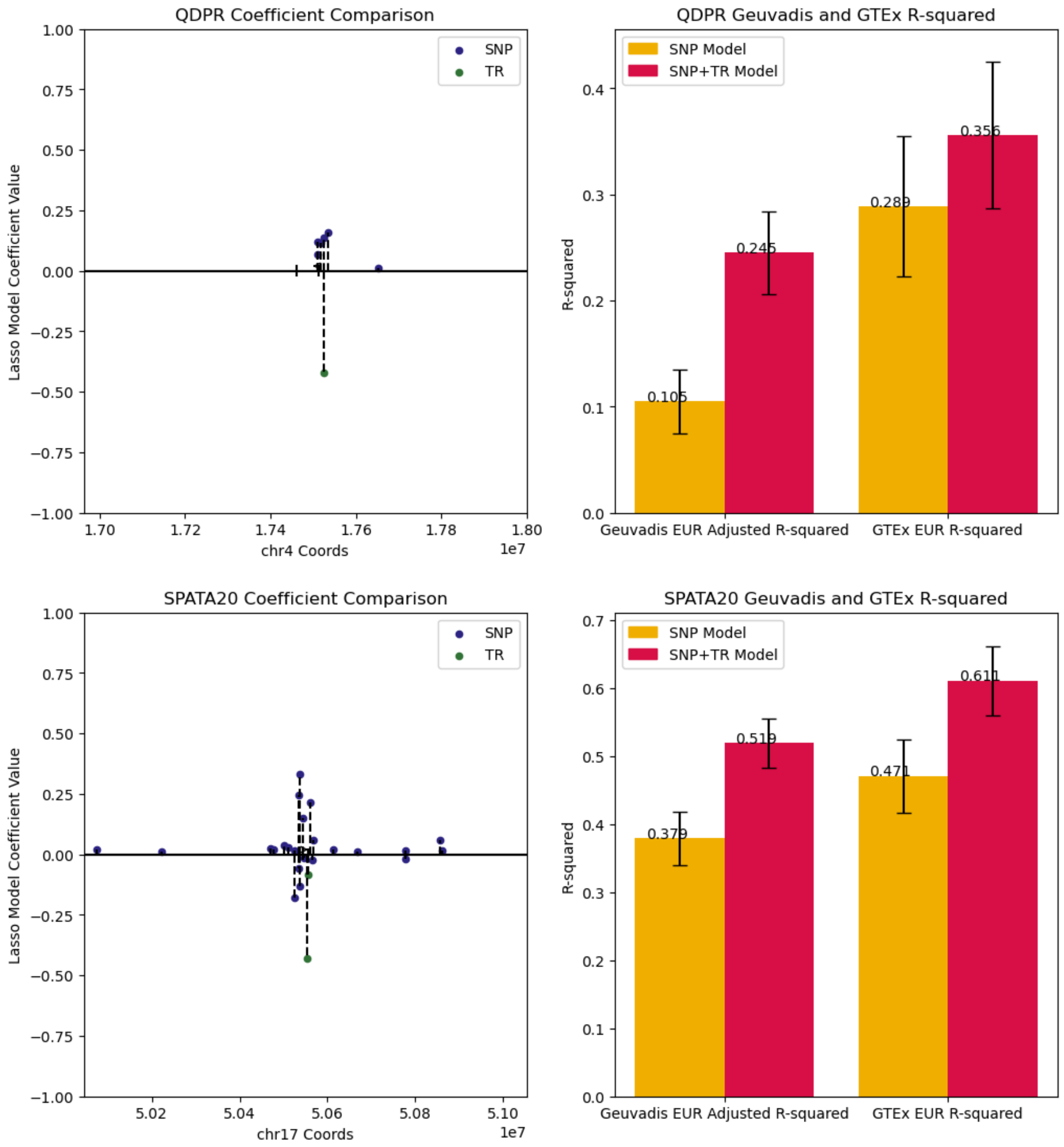
QDPR Coefficient Comparison
Lasso Model Coefficient Value
chr4 Coords
1e7
SNP
TR
QDPR Geuvadis and GTEx R-squared
SNP Model
SNP+TR Model
R-squared
0.105
0.245
0.289
0.356
Geuvadis EUR Adjusted R-squared
GTEx EUR R-squared
SPATA20 Coefficient Comparison
chr17 Coords
SPATA20 Geuvadis and GTEx R-squared
0.379
0.519
0.471
0.611

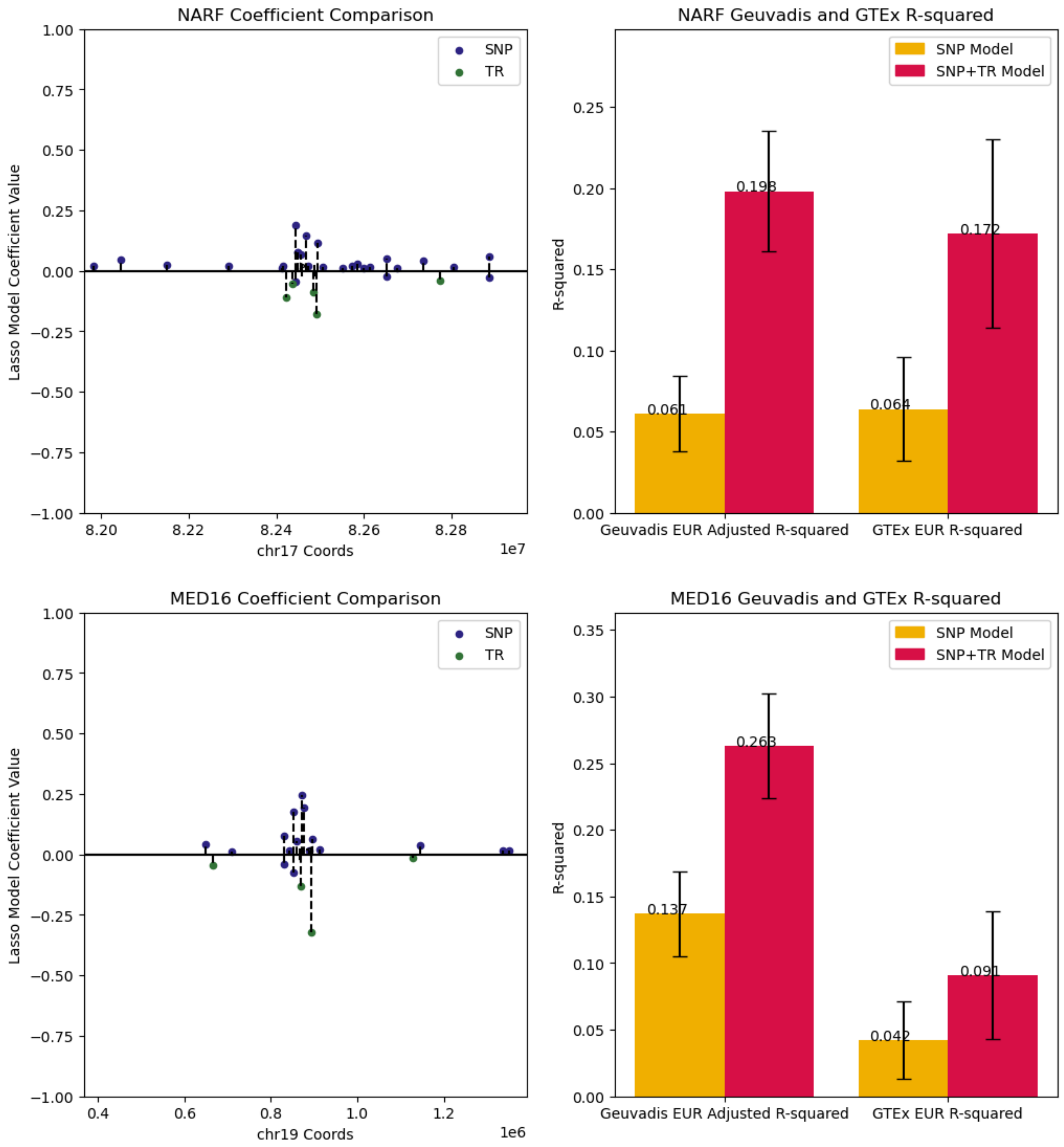
NARF Coefficient Comparison
Lasso Model Coefficient Value
chr17 Coords
1e7
SNP
TR
NARF Geuvadis and GTEx R-squared
R-squared
SNP Model
SNP+TR Model
0.061
0.198
0.064
0.172
Geuvadis EUR Adjusted R-squared
GTEx EUR R-squared
MED16 Coefficient Comparison
Lasso Model Coefficient Value
chr19 Coords
1e6
SNP
TR
MED16 Geuvadis and GTEx R-squared
R-squared
SNP Model
SNP+TR Model
0.137
0.263
0.042
0.091
Geuvadis EUR Adjusted R-squared
GTEx EUR R-squared

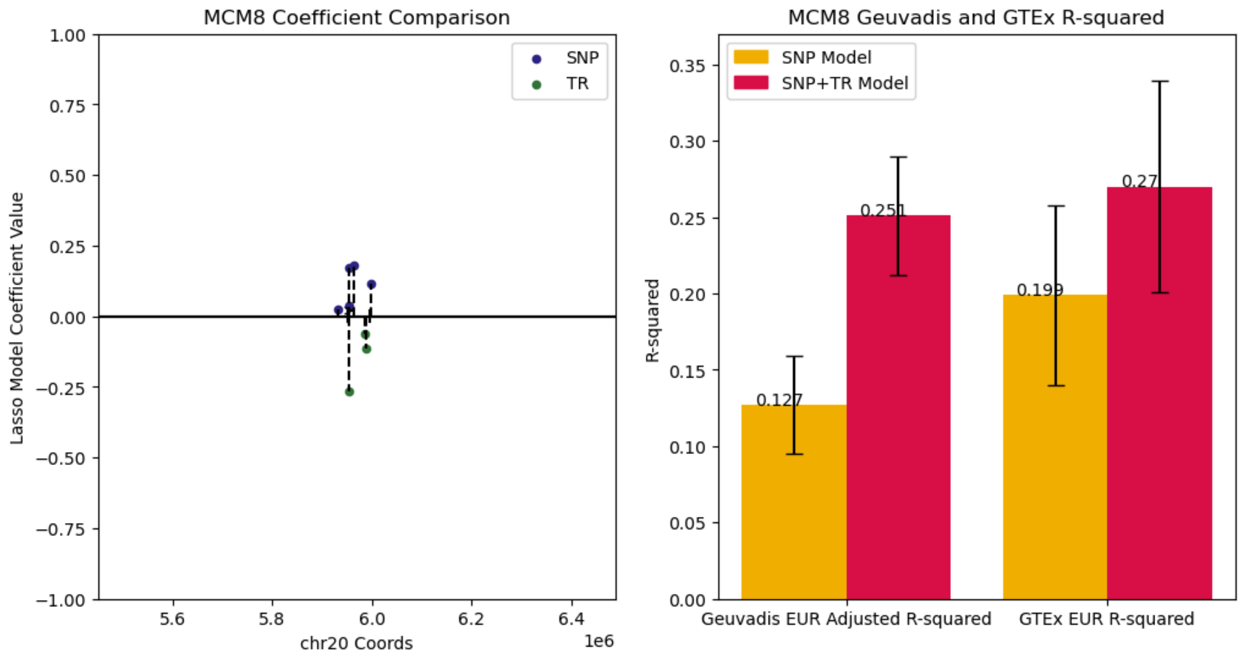


**Example genes for which incorporation of TRs show the strongest gains in gene expression prediction performance.** Data is shown for each of the genes in **Table 1** (besides *MDH1B*, which is shown in **Fig. 2c-d**) Left panels show the absolute value of model coefficients for SNPs (dark blue) and TRs (green) between the SNP-only models (upper y-axis; positive values) and SNP+TR models (lower y-axis; negative values). Right panels show the adjusted R-squared values for each gene in EUR Geuvadis samples (left bars) and the R-squared in European GTEx samples (right bars) for both the SNP only (yellow) and SNP+TR (red) models. Error bars give 95% confidence intervals. All genes except for *HLA-DPA1* had similar gain cross datasets.

#### Supplementary Figure 9

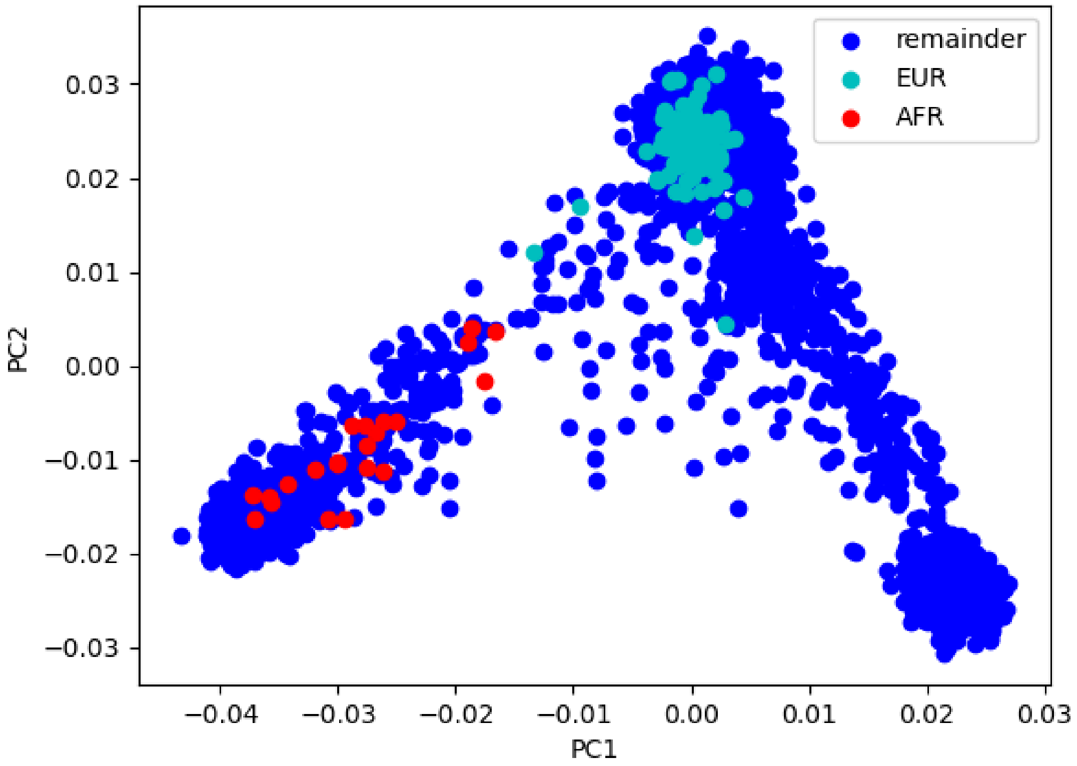


**Genotype principal component analysis (PCA) of the GTEx cohort.** This plot shows PC1 (x-axis) vs. PC2 (y-axis) for 3,184 samples from the GTEx (866) and 1000 Genomes (2,318) dataset. Samples highlighted in light blue and red are those for which expression data was available or EBV-transformed lymphocytes. We identified samples as European (EUR) as those with PC1 > -0.014 and PC2 > 0.01. The remaining samples with available expression data (red) were labeled as AFR based on their PC coordinates.

Supplementary Figure 10

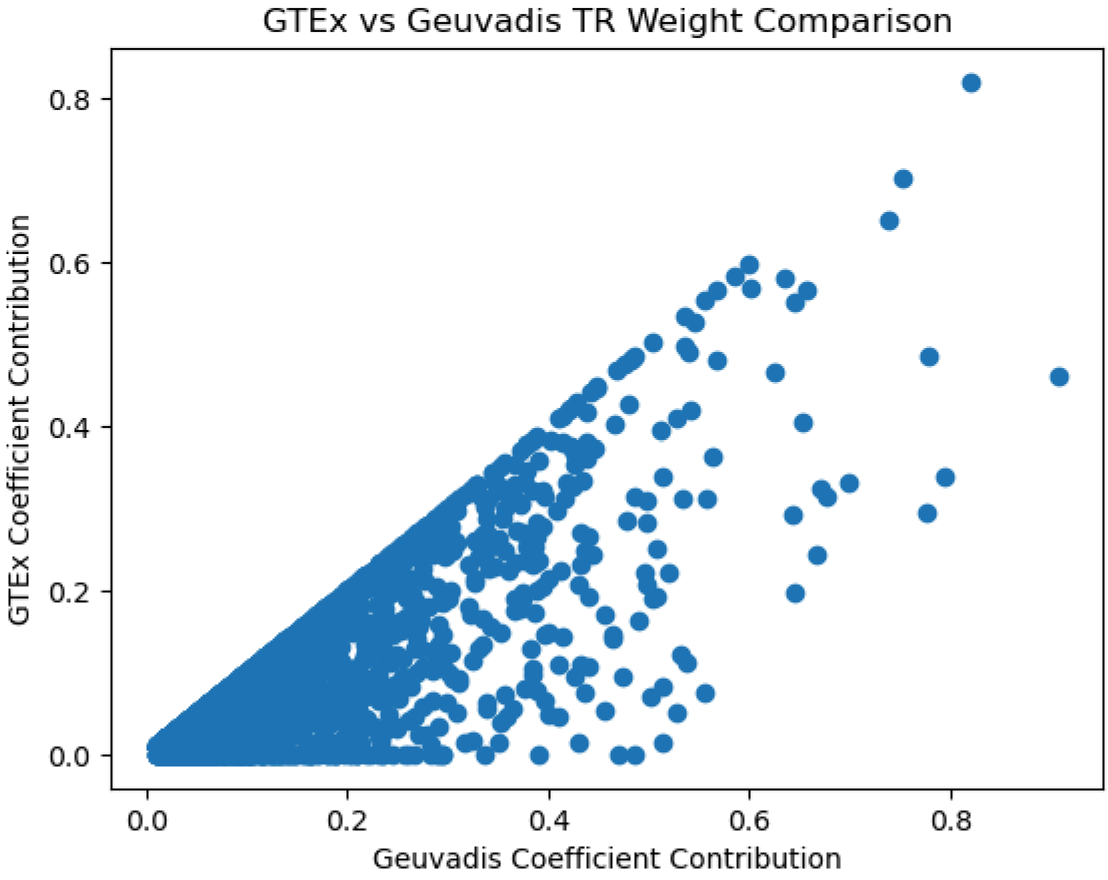


**Comparison of total TR model weights between Geuvadis and GTEx.** To generate TR genotypes for GTEx samples, we utilized imputation with the ensembleTR reference panel. This reference panel does not contain all TRs present in the Geuvadis TR genotype data and therefore there are TRs with non-zero coefficients that are not present in the GTEx data. This plot compares the ratio of the sum of the absolute value of TR coefficients where the TRs are present in the genotypes over the sum of the absolute value of all TR coefficients in the SNP+TR model. The ratio is compared between the Geuvadis dataset (x-axis) and the GTEx dataset (y-axis) for each gene that had at least one non-zero coefficient for a TR in the SNP+TR model (n=1,951).

### Supplementary Figure 11

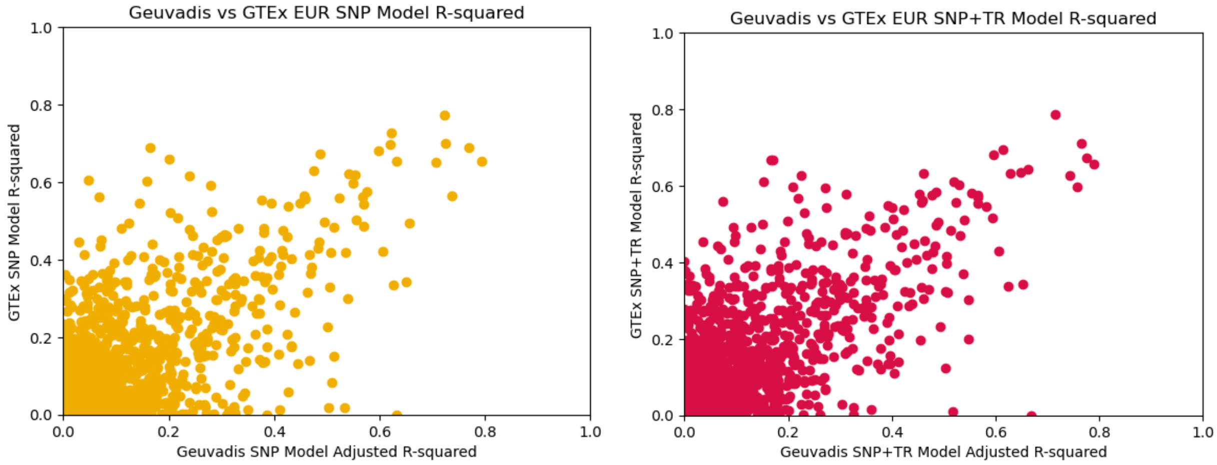


**Comparison of expression prediction model performance between Geuvadis and GTEx.** We compared the adjusted R-squared (Geuvadis, based on 3-fold cross-validation) to the R-squared (GTEx, using models trained in Geuvadis) for each gene in samples of European ancestry. The top panel shows data for SNP-only models (Pearson R=0.651; two-sided P=4.67e-185) and the bottom panel shows data for SNP+TR models (Pearson R=0.677; two-sided P=7.34e-206).

Supplementary Figure 12

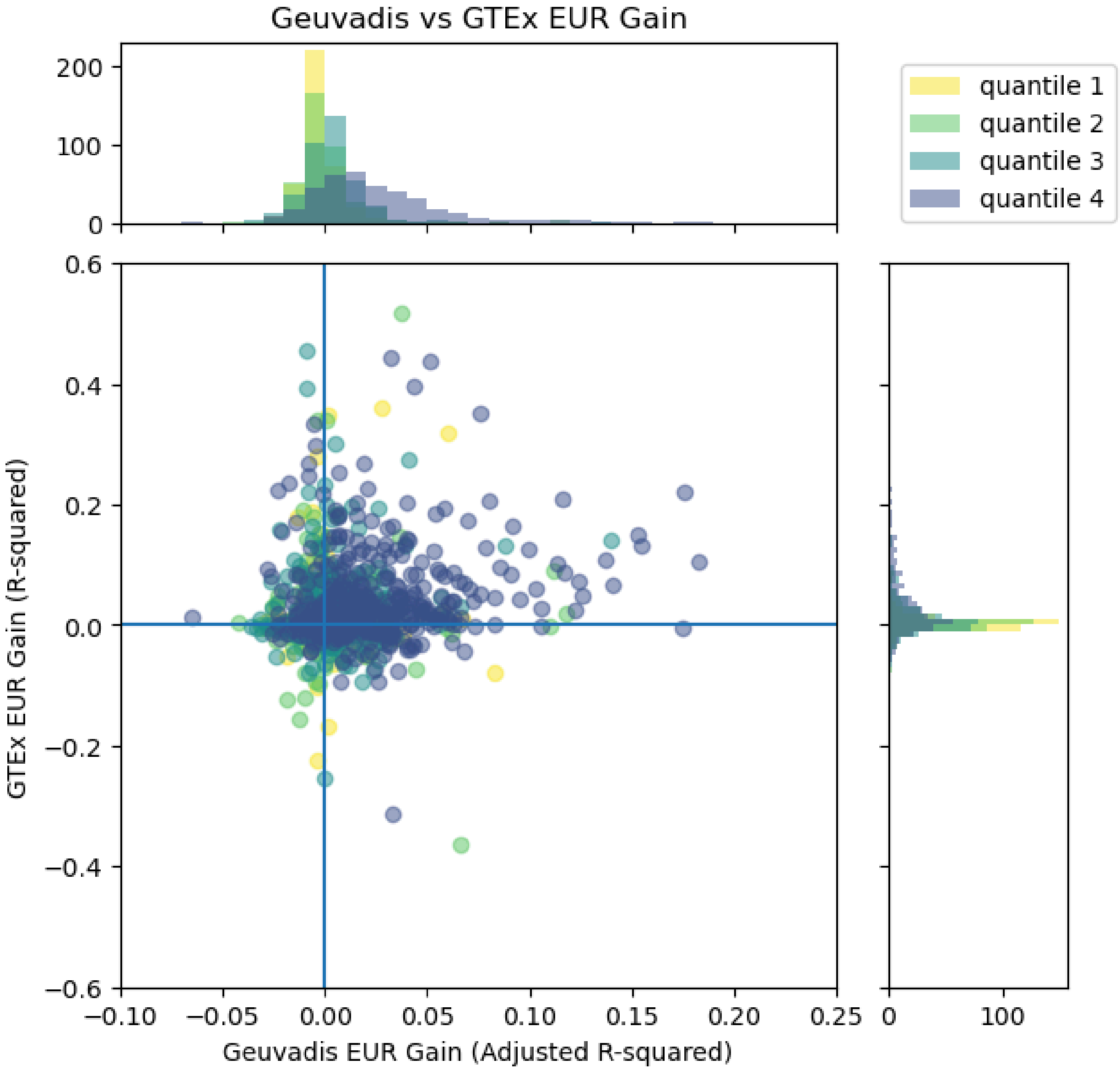


**Comparison of SNP+TR model gains across datasets.** We compared the gain in R-squared for SNP+TR vs. SNP-only models in the EUR cohort of Geuvadis (x-axis) vs. European samples in GTEx (y-axis) for each gene. Colors represent the quartile of each gene when ranked based on the total model weight assigned to TRs in trained models. Top and right panels show histograms of x-axis and y-axis values respectively stratified by these quartiles. Plot is the same as **Fig. 2e** except the y-axis is not truncated.

### Supplementary Figure 13

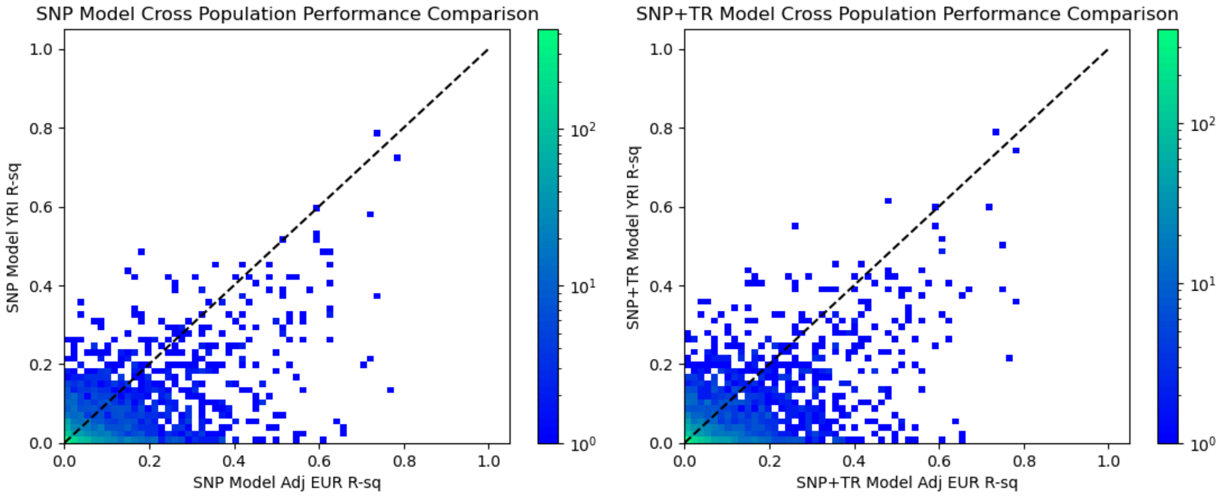


**Comparison of gene expression model performance in EUR vs. YRI samples in Geuvadis.** In each panel, the x-axis gives the adjusted R-squared based on EUR samples and the y-axis gives the R-squared for YRI samples in the Geuvadis cohort. The left panel shows SNP-only models (Pearson R=0.579, two-sided P=2.18e-228) and the right panel shows SNP+TR models (Pearson R=0.583, two-sided P=6.36e-225). The heatmap shows the density of data points (number of genes) at each coordinate.

### Supplementary Figure 14

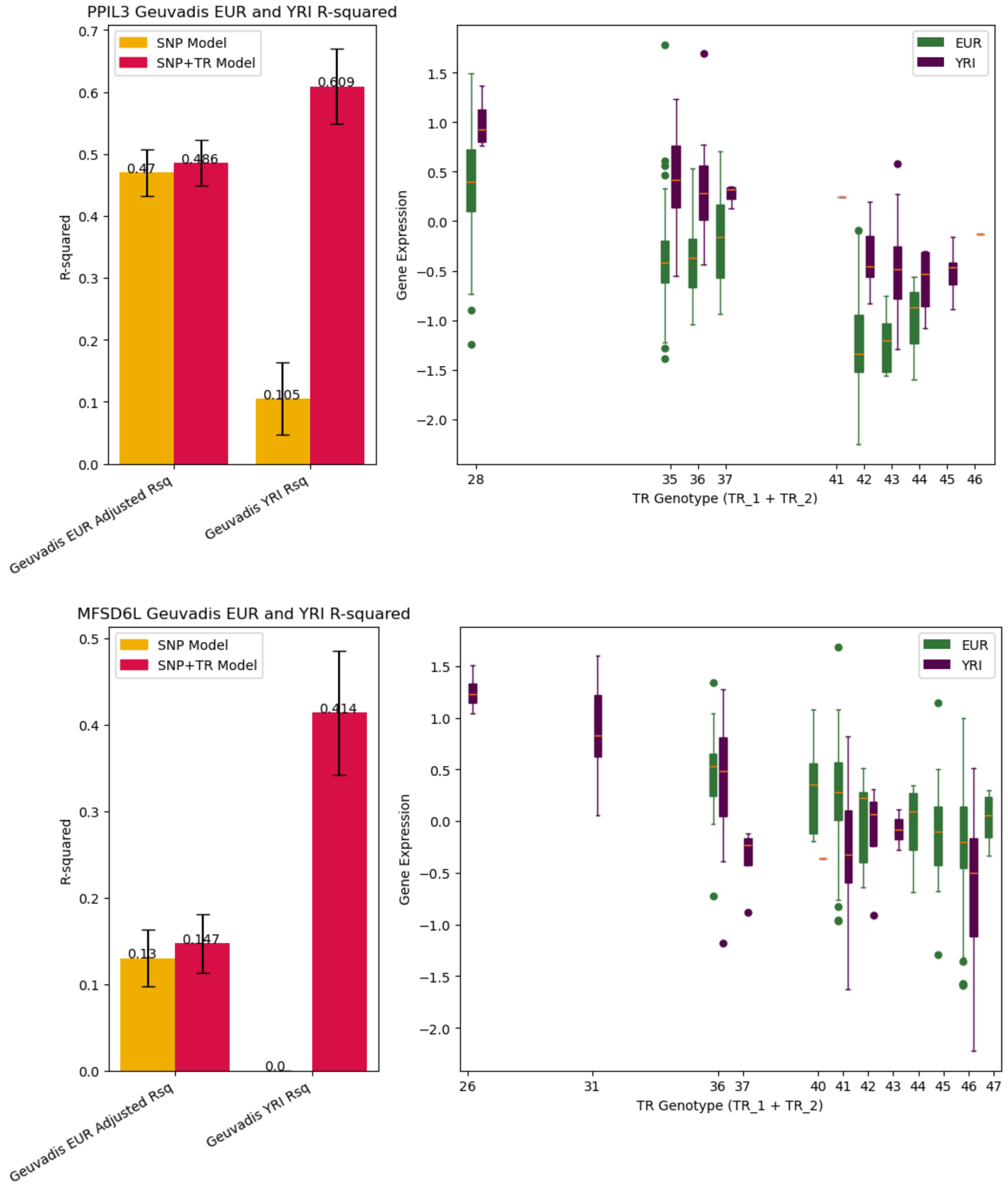

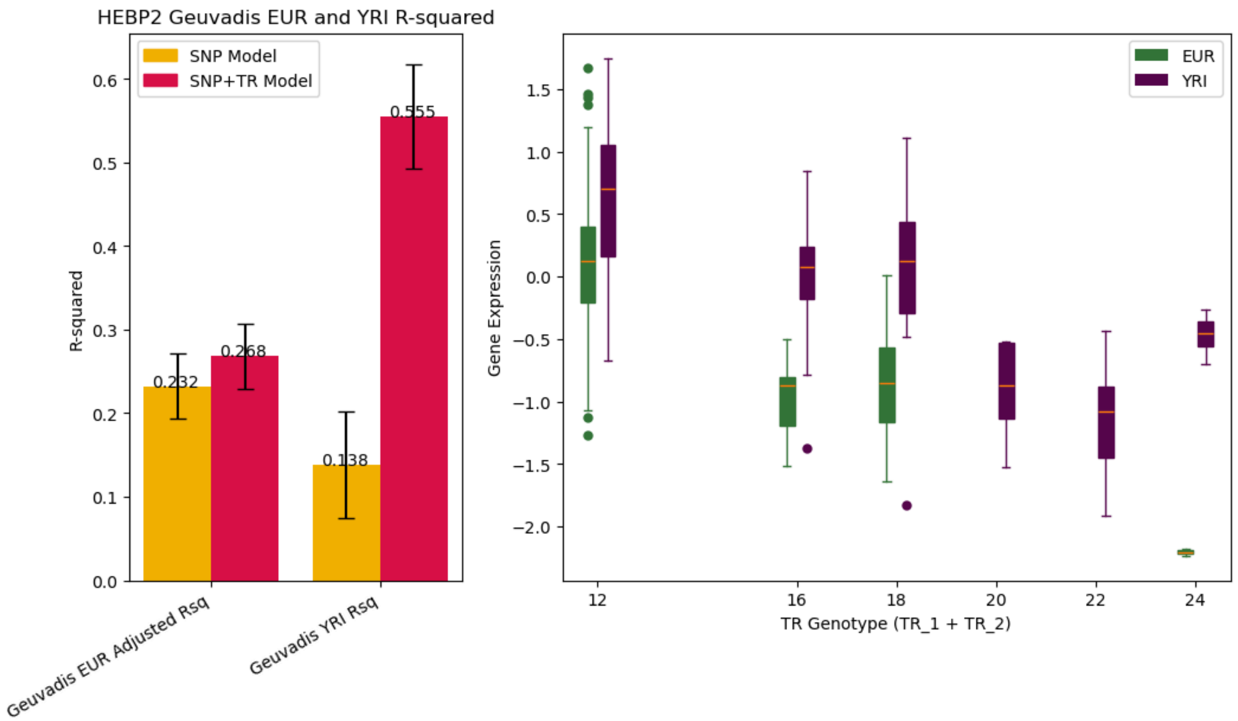
HEBP2 Geuvadis EUR and YRI R-squared
SNP Model
SNP+TR Model
R-squared
0.232
0.268
0.138
0.555
Geuvadis EUR Adjusted Rsq
Geuvadis YRI Rsq
EUR
YRI
Gene Expression
TR Genotype (TR_1 + TR_2)

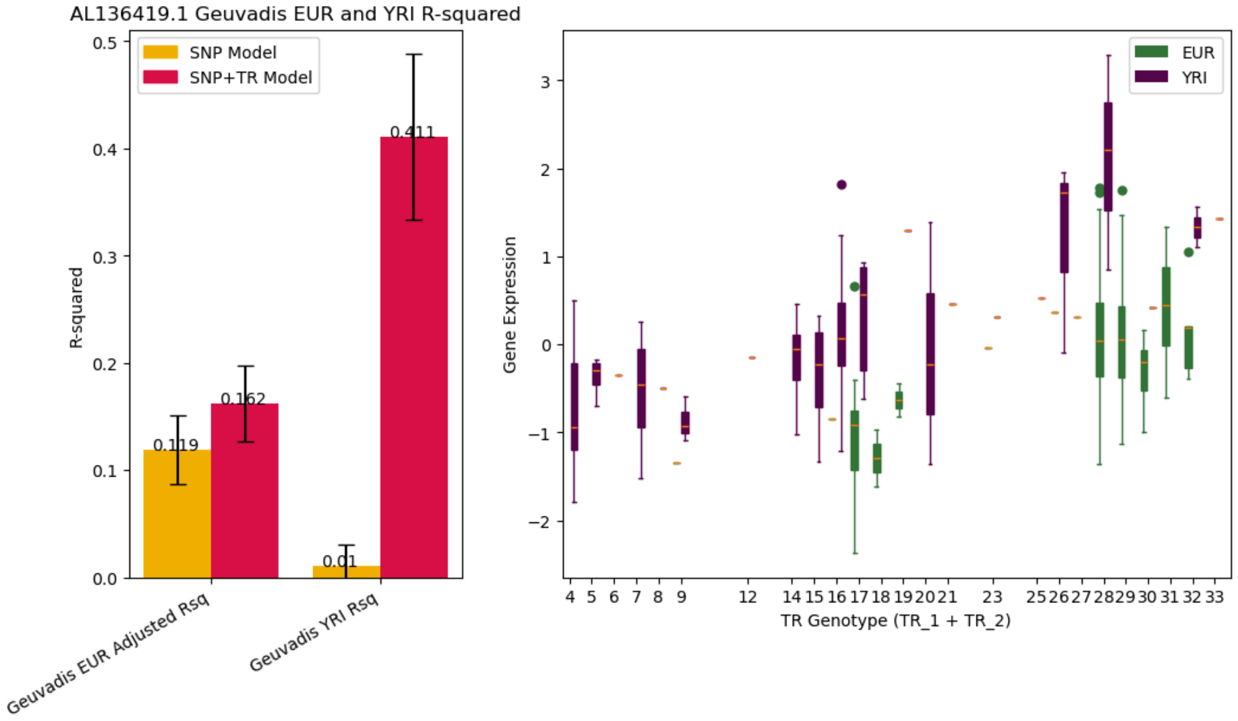
AL136419.1 Geuvadis EUR and YRI R-squared
SNP Model
SNP+TR Model
R-squared
0.119
0.162
0.01
0.411
Geuvadis EUR Adjusted Rsq
Geuvadis YRI Rsq
EUR
YRI
Gene Expression
TR Genotype (TR_1 + TR_2)

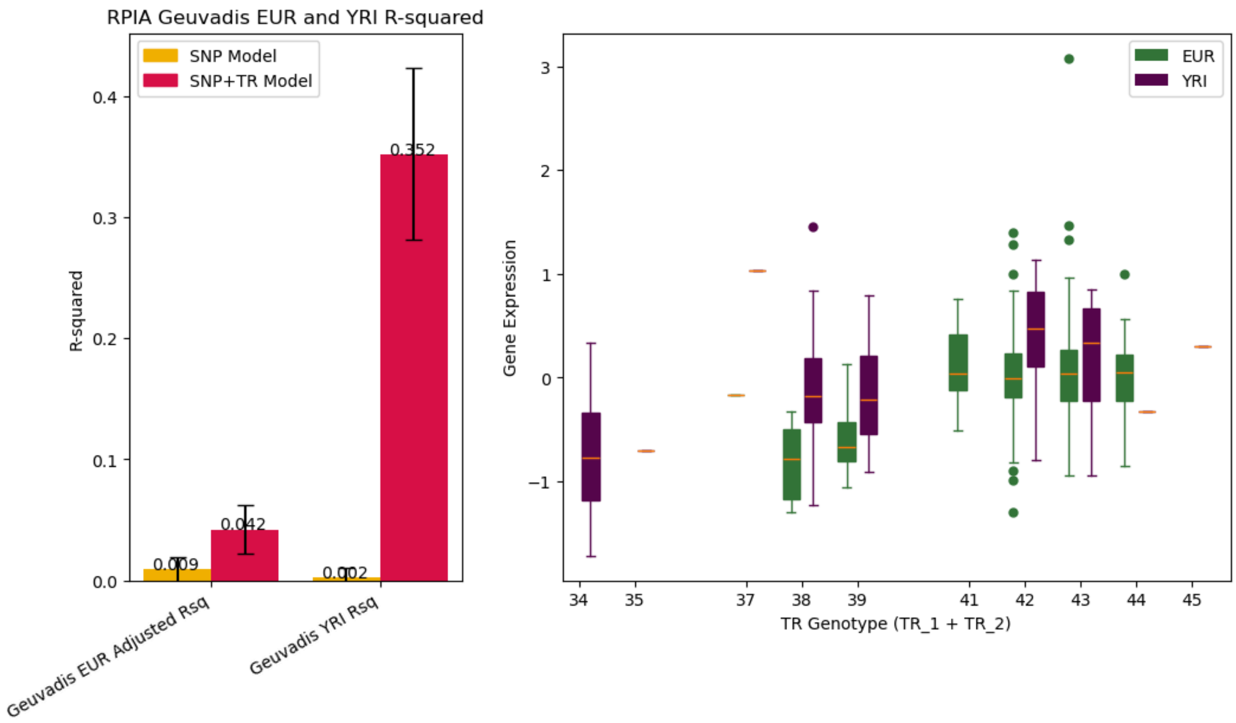
RPIA Geuvadis EUR and YRI R-squared
SNP Model
SNP+TR Model
R-squared
0.009
0.042
0.002
0.352
Geuvadis EUR Adjusted Rsq
Geuvadis YRI Rsq
EUR
YRI
Gene Expression
TR Genotype (TR_1 + TR_2)

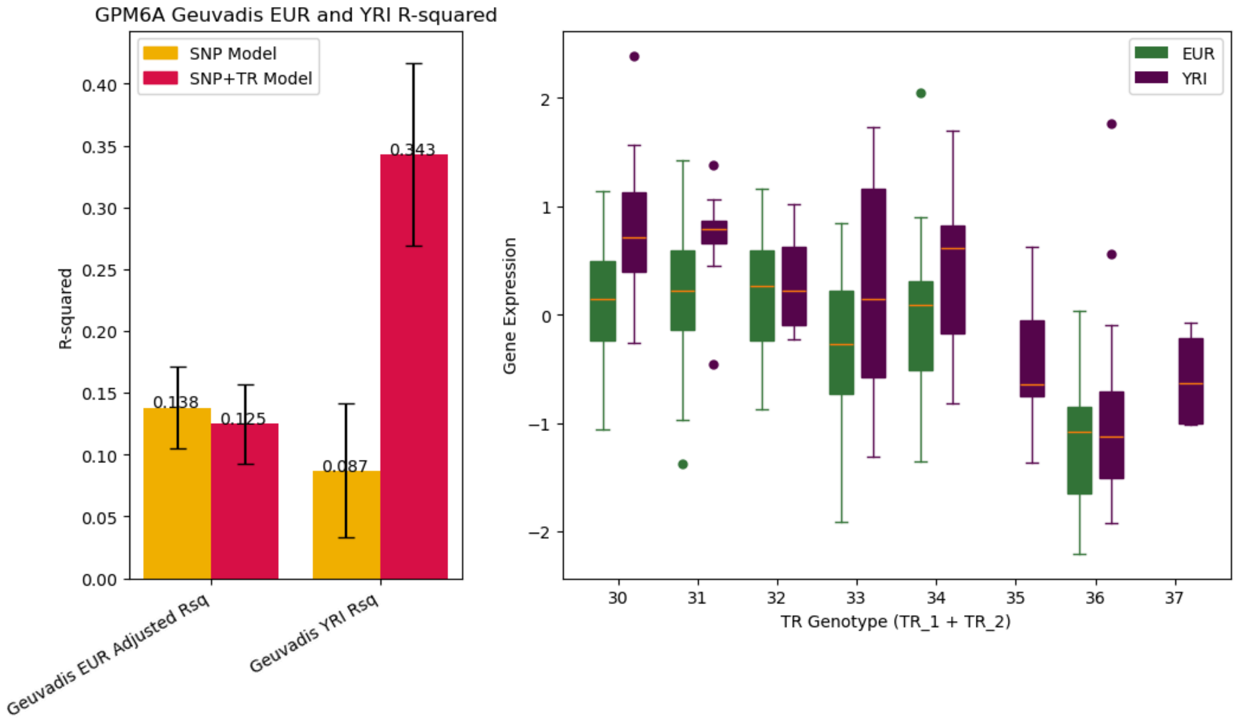
GPM6A Geuvadis EUR and YRI R-squared
SNP Model
SNP+TR Model
R-squared
0.138
0.125
0.087
0.343
Geuvadis EUR Adjusted Rsq
Geuvadis YRI Rsq
EUR
YRI
Gene Expression
TR Genotype (TR_1 + TR_2)

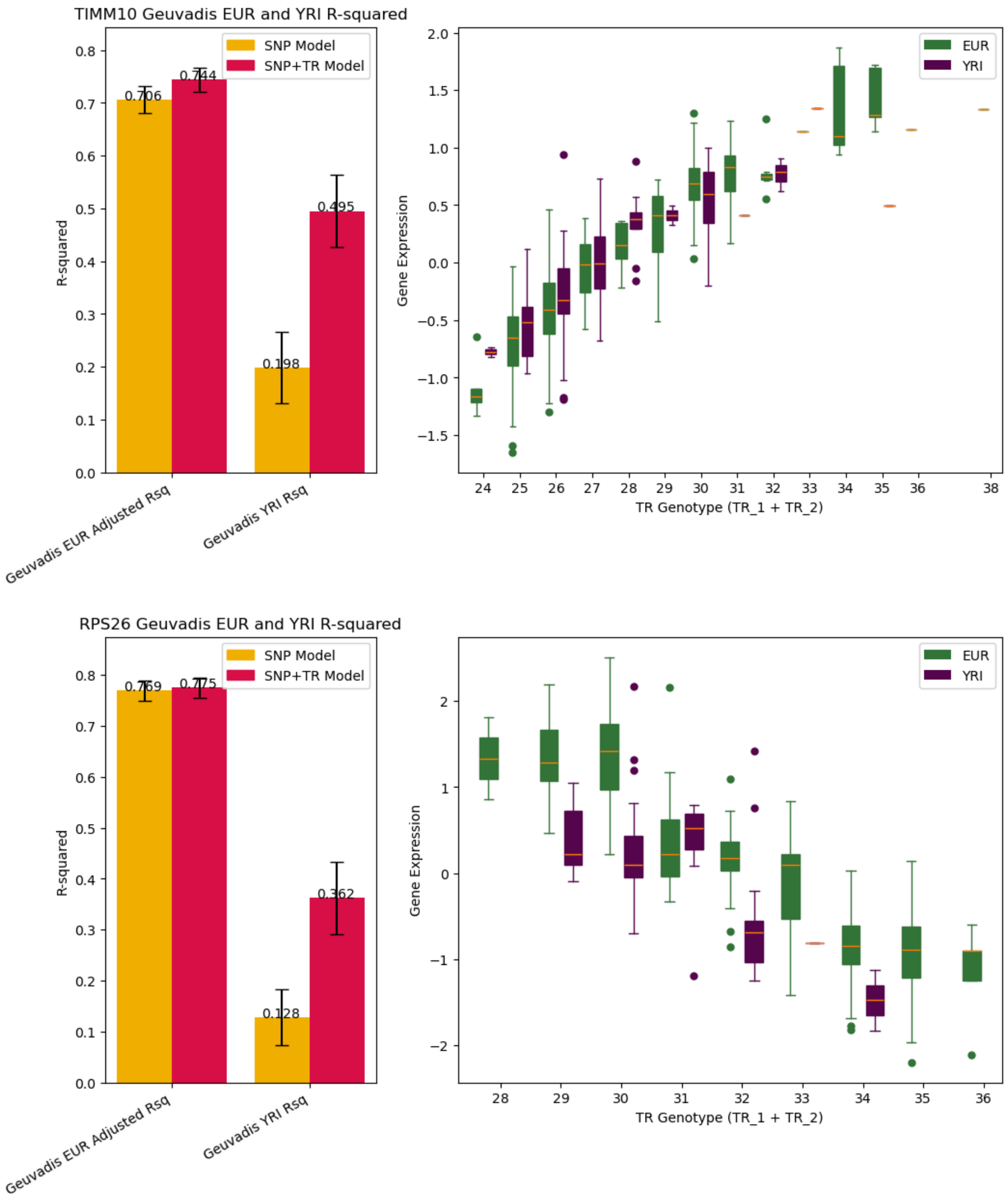

TIMM10 Geuvadis EUR and YRI R-squared
SNP Model
SNP+TR Model
R-squared
0.706
0.744
0.198
0.495
Geuvadis EUR Adjusted Rsq
Geuvadis YRI Rsq
EUR
YRI
Gene Expression
TR Genotype (TR_1 + TR_2)
RPS26 Geuvadis EUR and YRI R-squared
SNP Model
SNP+TR Model
R-squared
0.769
0.775
0.128
0.362
Geuvadis EUR Adjusted Rsq
Geuvadis YRI Rsq
EUR
YRI
Gene Expression
TR Genotype (TR_1 + TR_2)

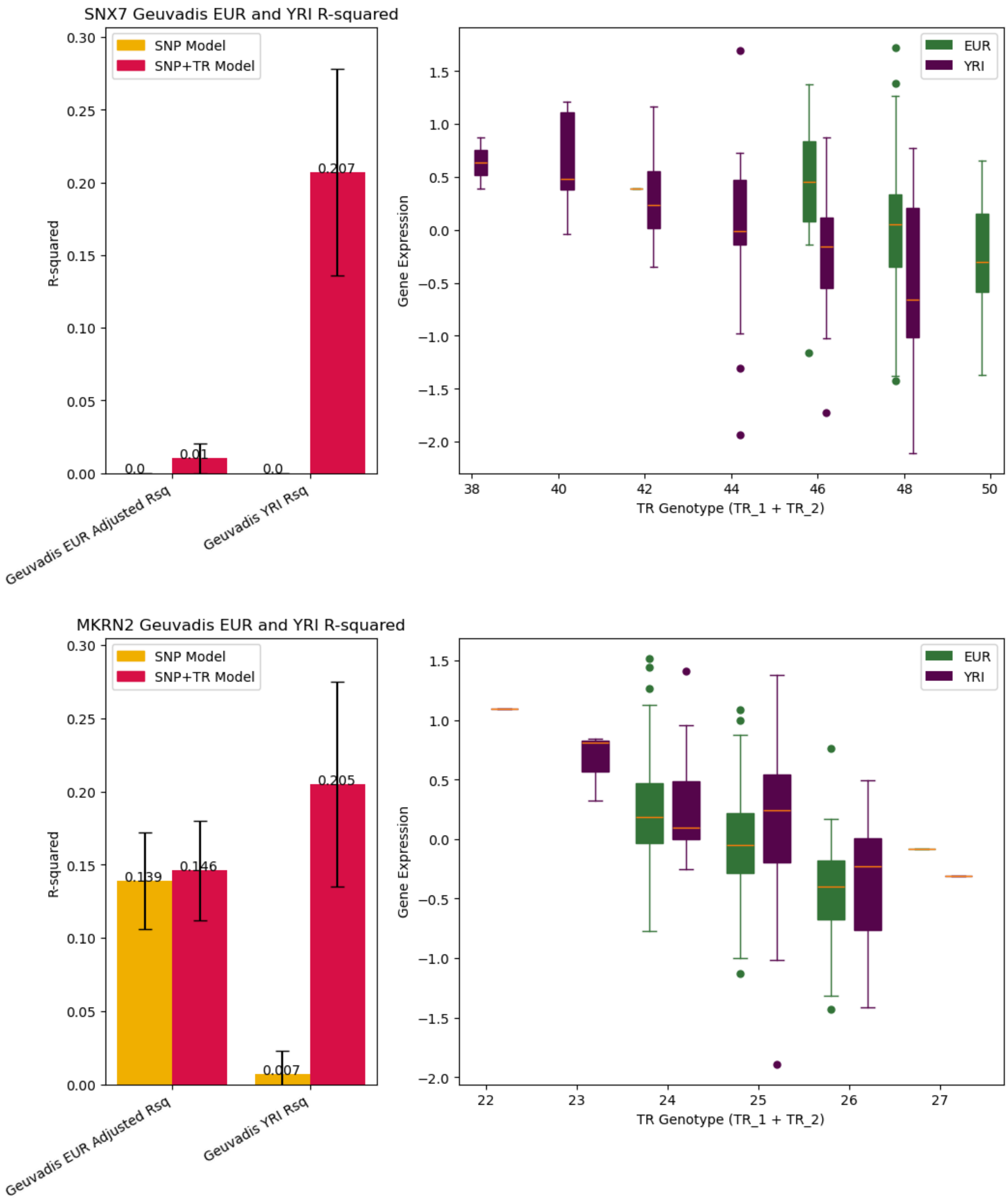


**Summary of gene expression models with the highest relative gains in YRI vs. EUR for SNP+TR - SNP-only models.** Left panels show the predictive performance in the Geuvadis EUR (left) and YRI (right) samples using models trained on European individuals. Gold=SNP-only models, red=SNP+TR models. Error bars give 95% confidence intervals. Right panels show associations between the TR with the largest coefficient for each SNP+TR model and expression of the gene. Plots show total repeat length (bp) across the two alleles of each individual (x-axis) vs. normalized gene expression in Geuvadis. Green=EUR samples and purple=YRI samples. Horizontal lines show median values, boxes span from the 25th percentile (Q1) to the 75th percentile (Q3), and whiskers extend to Q1-1.5*IQR (bottom) and Q3+1.5*IQR (top) where IQR is the interquartile range (Q3-Q1).

### Supplementary Figure 15

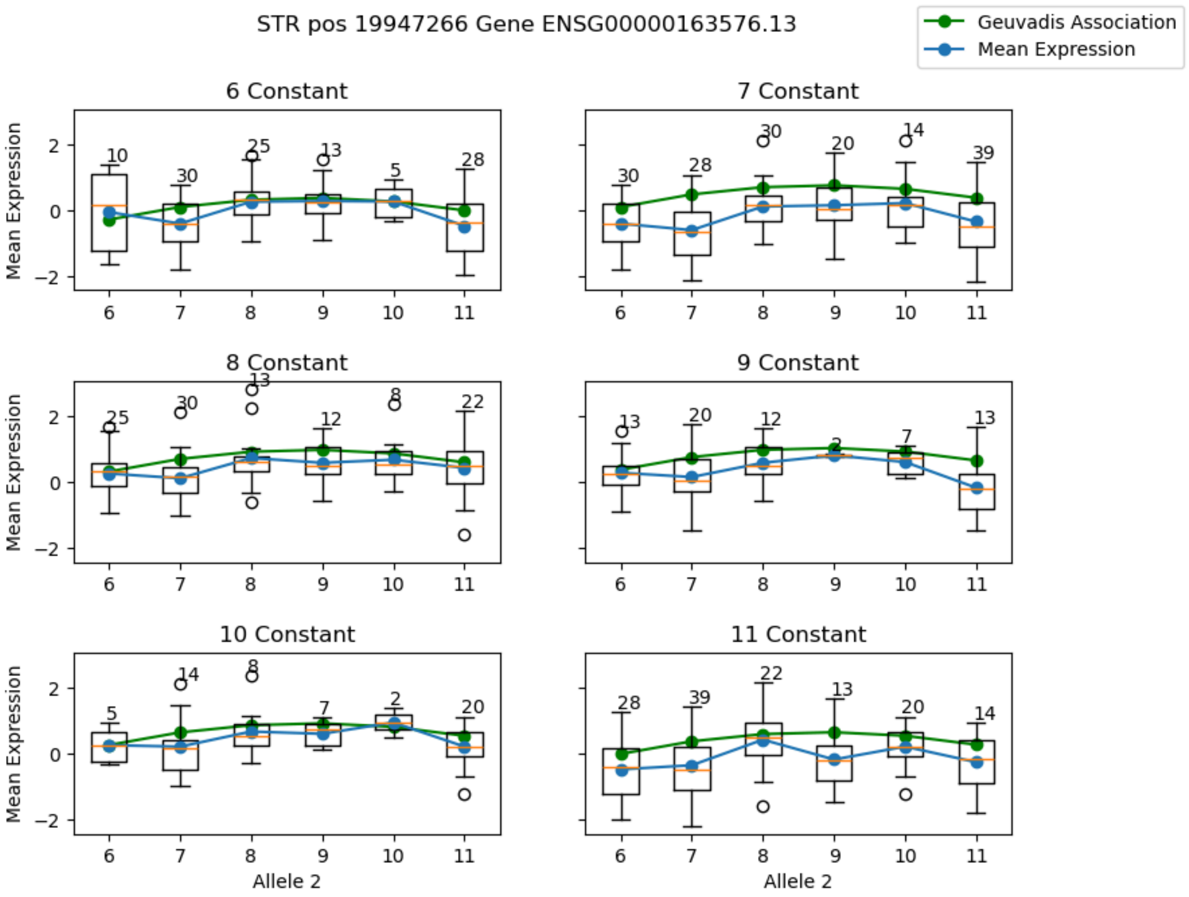


**Stratified associations with expression for a $CGG_n$ repeat non-linearly associated with *EFHB* expression.** Plots are the same as in **Fig. 4d**, except in each panel a different allele is held constant. For each panel, the x-axis gives the value of the allele not held constant in each individual. Blue=observed mean expression; green=trend learned from non-linear association testing. Each box is annotated with the number of samples that make up that distribution.

#### Supplementary Figure 16

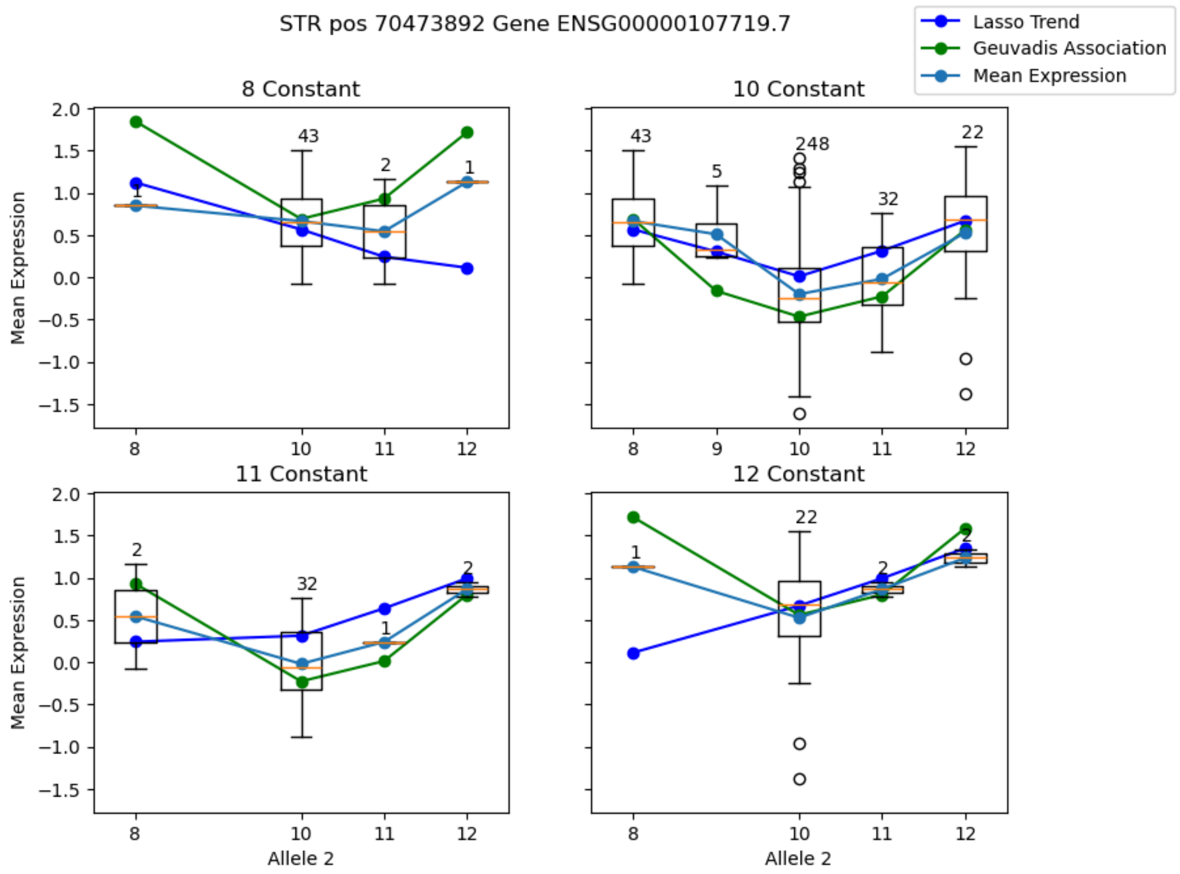


**Stratified associations with expression for a $C_n$ repeat non-linearly associated with *PALD1* expression.** Plots are the same as in **Fig. 4e**, except in each panel a different allele is held constant. For each panel, the x-axis gives the value of the allele not held constant in each individual. Blue=observed mean expression; green=trend learned from non-linear association testing. Each box is annotated with the number of samples that make up that distribution.

### Supplementary Table 2

| Enrichment Type | Odds Ratio | P-value | Num Pass Criteria |
|---|---|---|---|
| 1 | 1.72647059 | 1.10E-140 | 10269 |
| A | 1.70846098 | 2.47E-137 | 10209 |
| AAAT | 0.84494352 | 7.00E-03 | 261 |
| AAAAT | 0.72992245 | 1.85E-02 | 55 |
| AC | 0.72609279 | 2.09E-23 | 988 |
| AAC | 0.68579694 | 2.30E-03 | 63 |
| AAT | 0.66608993 | 1.56E-05 | 104 |
| 2 | 0.66261809 | 1.12E-51 | 1402 |
| AAAAG | 0.65356026 | 4.47E-03 | 43 |
| AT | 0.64260892 | 2.47E-14 | 271 |
| 5 | 0.64016508 | 3.92E-11 | 200 |
| AAAAC | 0.62501751 | 1.01E-04 | 63 |
| 3 | 0.6229998 | 1.55E-13 | 219 |
| AAAC | 0.60175455 | 4.27E-08 | 103 |
| 4 | 0.59459321 | 2.79E-36 | 534 |
| AG | 0.53471651 | 7.52E-16 | 140 |
| ATCC | 0.46684013 | 3.42E-03 | 13 |
| AAGG | 0.46012057 | 1.79E-06 | 31 |
| 6 | 0.4227857 | 3.59E-13 | 55 |
| AAAG | 0.41084145 | 7.18E-10 | 37 |
| AGAT | 0.31365499 | 2.88E-12 | 25 |

**Enrichment of repeat units at TRs with significant non-linear associations with gene expression.** For each unit sequence or length (bp), we performed a Fisher Exact Test to test for enrichment of each repeat unit in TRs with at least one significant non-linear association (ANOVA adjusted $P<0.05$) compared to all tested TRs. We only considered repeat units with at least 10 unique TRs passing ANOVA. Columns give the repeat unit sequence or length, odds ratio, two-sided P-value, and number of significant non-linear TRs.
